# Statistical and functional convergence of common and rare variant risk for autism spectrum disorders at chromosome 16p

**DOI:** 10.1101/2022.03.23.22272826

**Authors:** Daniel J. Weiner, Emi Ling, Serkan Erdin, Derek J.C. Tai, Rachita Yadav, Jakob Grove, Jack M. Fu, Ajay Nadig, Caitlin E. Carey, Nikolas Baya, Jonas Bybjerg-Grauholm, iPSYCH Consortium, ASD Working Group of the Psychiatric Genomics Consortium, ADHD Working Group of the Psychiatric Genomics Consortium, Sabina Berretta, Evan Z. Macosko, Jonathan Sebat, Luke J. O’Connor, David M. Hougaard, Anders D. Børglum, Michael E. Talkowski, Steve A. McCarroll, Elise B. Robinson

## Abstract

The dominant human genetics paradigm for converting association to mechanism (“variant-to-function”) involves iteratively mapping individual associations to specific SNPs and to the proximal genes through which they act. In contrast, here we demonstrate the feasibility of extracting biological insight from a very large (>10Mb) region of the genome, and leverage this approach to derive insight into autism spectrum disorder (ASD). Using a novel statistical framework applied in an unbiased scan of the genome, we identified the 33Mb p-arm of chromosome 16 (16p) as harboring the greatest excess of common polygenic risk for ASD. This region includes the recurrent 16p11.2 copy number variant (CNV) – one of the largest single genetic risk factors for ASD, and whose pathogenic mechanisms are undefined. Analysis of bulk and single-cell RNA-sequencing data from post-mortem human brain samples revealed that common polygenic risk for ASD within 16p associated with decreased average expression of genes throughout this 33-Mb region. Similarly, analysis of isogenic neuronal cell lines with CRISPR/Cas9-mediated deletion of 16p11.2 revealed that the deletion also associated with depressed average gene expression across 16p. The effects of the rare deletion and diffuse common variation were correlated at the level of individual genes. Finally, analysis of chromatin contact patterns by Hi-C revealed patterns which may explain this transcriptional convergence, including elevated contact throughout 16p, and between 16p11.2 and a distal region on 16p (Mb 0-5.2) which showed the greatest gene expression changes in both the common and rare variant analyses. These results demonstrate that elevated 3D chromatin contact may coordinate genetic and transcriptional disease liability across large genomic regions, exemplifying a novel approach for extracting biological insight from genetic association data. As applied to ASD, our analyses highlight the 33Mb p-arm of chromosome 16 as a novel locus for ASD liability and provide insight into disease liability originating from the 16p11.2 CNV.

## Introduction

Genome-wide association studies have productively identified robust statistical associations between thousands of common genetic variants and traits.^1^ However, most associations are non-coding, complicating efforts to identify the genes that mediate these associations.^2,3^ A dominant approach is to fine-map associations to individual variants and then to their nearby target genes.^4,5^ While there are numerous examples of success,^5,6^ functional interpretation of individual genetic variants remains a critical bottleneck. Moreover, most complex trait heritability often does not reside with these individually significant associations, but is rather scattered across thousands of individually non-significant loci across the genome.^7^

Autism spectrum disorder (ASD) provides a compelling example of the need to jointly interpret many classes of genetic variation.^8–14^ While common polygenic variation is the largest genetic risk factor for ASD on a population level, extracting biological insight from this predominantly non-coding signal is challenging.^11^ Similarly, *de novo* recurrent copy number variants (CNVs), which are strongly associated with ASD, often encompass many genes with generally undefined downstream mechanisms.^8,13,15–17^ For example, while deletion of the 0.7Mb, 31-gene locus at chromosome 16p11.2 is among the most common and largest single genetic risk factors for ASD^8,18^, exactly how the deletion confers ASD liability has remained undetermined despite considerable inquiry.^19–23^ Thus, a critical open question is whether regional polygenic signals colocalize with recurrent large CNVs, and whether this colocalization can highlight uncommonly relevant areas of the genome for ASD risk. In particular, given that both regions of polygenic risk and recurrent large CNVs span many genes and influence chromatin structure and gene regulatory landscapes,^24–27^ large chromatin landscapes have the potential to unify analysis of regional polygenic and rare variation.

In order to examine polygenic risk arising from regions of the genome, including regions harboring ASD-associated CNVs, we developed the stratified polygenic transmission disequilibrium test (S-pTDT), which extends the trio-based polygenic transmission disequilibrium test (pTDT) to specific genomic annotations. Using S-pTDT and 9,383 European-ancestry ASD trios, we performed an unbiased genome-wide search for excess over-transmission of ASD polygenic risk. Unexpectedly, the greatest excess localized to the 33Mb p-arm of chromosome 16 (16p), the region which includes the recurrent, ASD-associated and mechanically cryptic 16p11.2 CNV. Further linking the 16p11.2 CNV with the broader p-arm of the chromosome, *in vitro* deletion of the 16p11.2 locus was associated with decreased average expression of 200 neuronally expressed genes on chromosome 16p. Similarly, an increased ASD polygenic risk score (PRS) constructed exclusively with 16p variants was associated with decreased 16p mean brain gene expression across multiple cohorts with paired genotype and brain gene expression measurements. These transcriptional effects of the 16p11.2 CNV and 16p ASD PRS were correlated at the level of individual gene expression on 16p, suggesting mechanistic convergence of common and rare variant ASD liability in the region. We observed chromatin contact patterns which we hypothesize explain this transcriptional convergence: uncommonly high within-16p chromatin contact in two independent Hi-C datasets and b) increased contact between the 16p11.2 CNV and a distal region on 16p (Mb 0-5.2) with the greatest observed gene expression changes. Our results motivate a model of convergent common and rare genetic risk factors for ASD at 16p, and more broadly suggest that elevated 3D chromatin contact may facilitate coordinated genetic and transcriptional disease liability within very large regions of the genome.

## Results

### Stratified-pTDT estimates polygenic overtransmission of ASD PRS at 16p

Children with ASD inherit more polygenic risk for ASD from their unaffected parents than expected by chance (“overtransmission”).^12^ Using pTDT, we observe mean overtransmission of ASD PRS within European ancestry trios from three different ASD trio cohorts: the Psychiatric Genomics Consortium (PGC)^11^ (n = 4,335 trios, 0.20 SD overtransmission, P = 1.53e-37), the Simons Simplex Collection (SSC)^28^ (n = 1,851 trios, 0.19 SD overtransmission, P = 1.29e-17), and Simons Foundation Powering Autism Research (SPARK)^29^ (n = 3,197 trios, 0.17 SD overtransmission, P = 6.38e-21), all using a PRS generated from an external GWAS from the Danish iPSYCH collection (19,870 cases, 39,078 controls) (Supplementary Figures 1-2). As biological insights from ASD’s common variant risk factors have been limited, we aimed to leverage the statistical power of pTDT to identify regions of the genome of excess common variant relevance in ASD. Thus, we introduce stratified-pTDT (S-pTDT), which estimates transmission in parent-child trios of PRS constructed from small sets of SNPs (**Figure 1A**). Like pTDT, S-pTDT is a within family test, which prevents spurious association due to population stratification and many types of ascertainment bias.^12^

**Figure 1:**
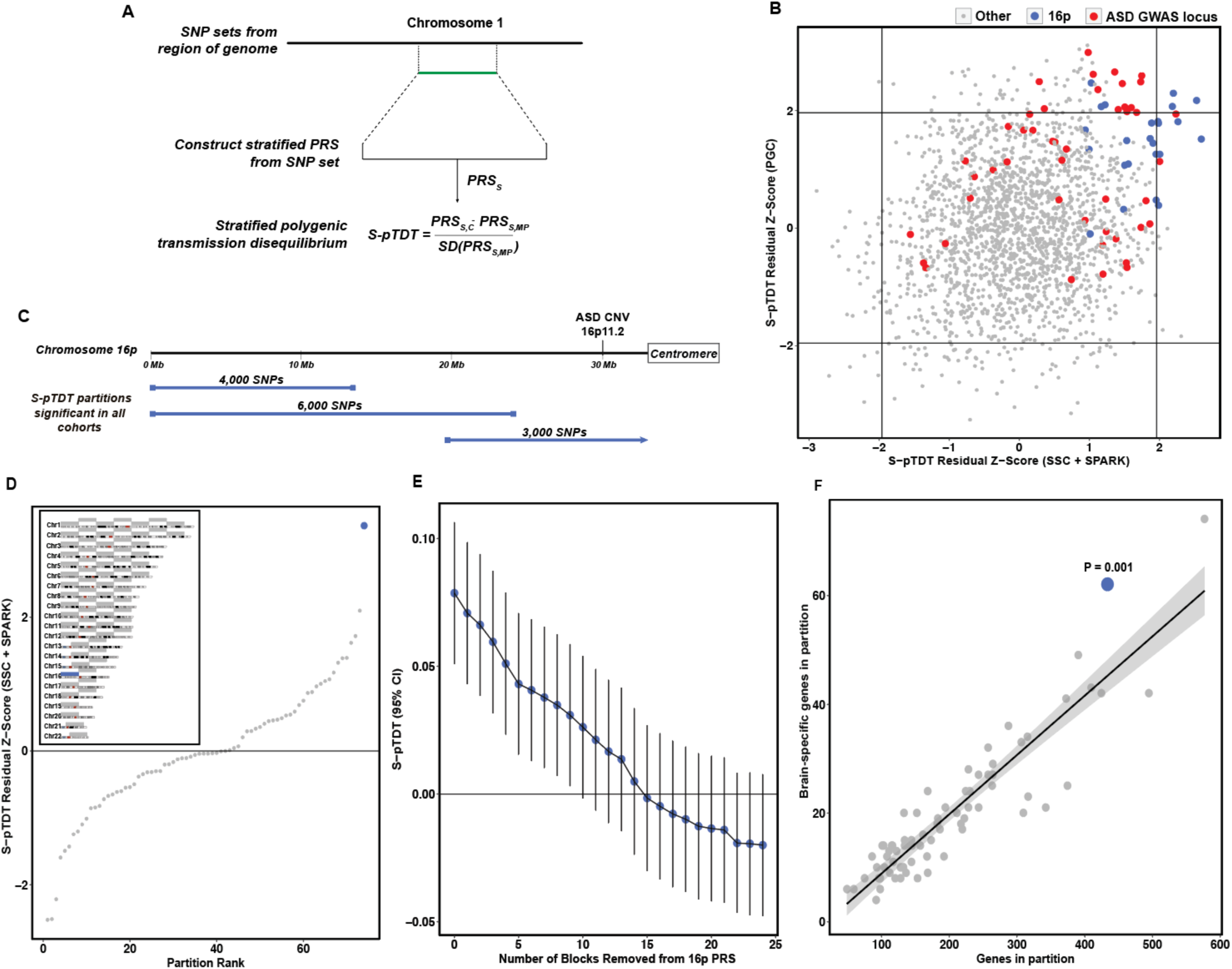
Stratified polygenic transmission disequilibrium localizes regional ASD polygenic overtransmission at 16p. **(A)** S-pTDT estimates transmission of stratified polygenic scores from parents to their children. Stratified polygenic scores are constructed from a continuous block of SNPs, denoted in green (though in general, S-pTDT can handle stratified PRS constructed from the union of multiple SNP sets). The S-pTDT value for a parent-child trio is the difference between the stratified polygenic score in the child and the expectation formed by the average stratified polygenic score of their parents, normalized by the standard deviation of the mid-parent stratified polygenic score for the group of trios. **(B)** European ancestry ASD probands in the combined Simons Simplex Collection (SSC) and Simons Foundation Powering Autism Research (SPARK) cohorts (x-axis, n = 5,048 trios) and Psychiatric Genomics Consortium (y-axis, n = 4,335) over-inherit stratified polygenic scores constructed from partitions on the p-arm of chromosome 16 (16p, blue dots) and partitions with ASD GWAS loci (red dots) using a non-overlapping ASD GWAS from the Danish iPSYCH collection (n = 19,870 cases, 39,078 controls). The axes are in units of S-pTDT residual z-scores from a model regressing out SNP number and partition size in base pairs from the partition S-pTDT estimate. Horizontal and vertical lines are positioned at z-scores +/-1.96. **(C)** The three partitions that are nominally significant in both combined SSC + SPARK and PGC (blue bars) collectively span 16p. **(D)** Inset: The 16p partition (blue bar) is compared with 73 other 33Mb partitions (gray bars) spanning the genome. Main panel: ASD S-pTDT analysis of the combined SSC and SPARK cohorts with stratified polygenic scores constructed from the 33Mb partitions. Each dot shows the S-pTDT value for the partition in units of residual z-score; the blue dot is 16p. **(E)** SSC + SPARK ASD S-pTDT decays gradually with successive removal of most associated remaining LD-independent block on 16p. The y-axis is the S-pTDT estimate (+/-95% CI) for transmission of a stratified polygenic scores constructed from the union of remaining blocks. The S-pTDT estimates for each of the LD-independent blocks are shown in Supplementary Figure 8. **(F)** 16p is gene dense and enriched in brain-specific genes. Each point is a 33Mb partition as defined in D, and 16p is shown in blue. Brain-specific genes are defined as genes in the top 10% of specific expression in cortex relative to non-brain tissues in GTEx (Finucane 2018). P-value is derived from the residual z-score in a linear model regressing the y on x variable, and the shaded region is a 95% CI.

We first asked whether S-pTDT could identify any regions of the genome with transmission of ASD polygenic risk significantly over or under genome-wide expectation for a region of that size. To do this, we constructed stratified PRS from adjacent blocks of SNPs, yielding 2,006 (often overlapping) partitions collectively covering the whole genome (median number of SNPs per block: 3,000, minimum length: 4.3Mb, maximum length: 52.9Mb, median length: 11.7Mb, Supplementary Figure 3, Methods). We then performed S-pTDT on each partition, first estimating transmission in the 5,048 trios from SSC+SPARK, then estimating transmission in the 4,335 trios from PGC. As expected given the robust genome-wide pTDT overtransmission, most of these stratified partitions have a point estimate of overtransmission, and the degree of overtransmission increases with number of SNPs in the partition and size of the partition (Supplementary Figure 4). In order to estimate the extent to which transmission of each region differed from genome-wide expectation, we constructed a linear model, regressing S-pTDT transmission on the number of SNPs in the partition and the length of the partition (Methods). This model yields a residual z-score for each partition, which estimates, in standard deviations, how much more or less transmission there is than there is expected relative to genome-wide patterns.

Transmission of regional polygenic risk for ASD is correlated between SSC/SPARK and PGC trios (r = 0.21 (P = 3.4e-21), **Figure 1B**, Supplementary Figure 5), which indicates stability in the S-pTDT rankings despite each partition including on average only 0.1% of SNPs, the vast majority of partitions containing no ASD GWAS loci, and phenotypic/genetic heterogeneity among the iPSYCH ASD GWAS and ASD trio cohorts. Partitions that include ASD GWAS loci^11^ are enriched among positive (z-score > 0) S-pTDT scores in both SSC/SPARK and PGC: 29/46 (63%) GWAS loci are top right quadrant, compared with expectation of 12 partitions (chi-sq P-value = 1.68e-8) (**Figure 1B**, red points). This observation is consistent with the expectation that ASD cases on average over-inherit ASD risk alleles.

Unexpectedly, partitions with large S-pTDT z-scores cluster on the p-arm of chromosome 16 (from here, 16p, approximately 0-33Mb) (**Figure 1B**, blue points): of the 12 partitions with the largest average S-pTDT z-score across SSC/SPARK and PGC, 5 are on 16p, while the other 7 localize to ASD GWAS loci. The three partitions that are nominally enriched in both data sets (S-pTDT z-score > 1.96) collectively tile the entirety of 16p (**Figure 1C**). Given that the highly over-transmitted regions span the p-arm of chromosome 16, we constructed a new partition spanning the p-arm (33Mb), and compared it to the 73 other, non-overlapping 33Mb regions found in the human genome (**Figure 1D inset**). The excess overtransmission at 16p becomes even more apparent in this analysis, with an S-pTDT z-score in SSC+SPARK of 3.37 (**Figure 1D**). In contrast, the same common variants at 16p are not over-transmitted to 1,509 unaffected siblings in SSC (S-pTDT z-score = -0.06 (P = 0.95), Supplementary Figure 6).

While 16p does not contain a genome-wide significant locus for ASD (Supplementary Figure 7), we nevertheless sought to determine whether the S-pTDT signal at 16p could be explained by one or a small number of common variant associations. We partitioned the 16p region into 25 adjacent blocks with low between-block LD (median length: 1.31 Mb) and assessed S-pTDT signal for each.^30^ Consistent with absence of a single driving locus in the region, the association signal was diffuse (Supplementary Figure 8) and decayed gradually with successive removal of the most over-transmitted blocks (**Figure 1E**, Methods). Restated, the S-pTDT association at 16p does not appear to be driven by a single coding or regulatory locus in the region, but exists more diffusely across the 33Mb segment of genome.

We performed a number of additional analyses to further interrogate the S-pTDT finding at 16p. First, ASD cases with a neurodevelopmental-disorder-associated CNV on 16p (1.0% of cases in SSC/SPARK) did not drive the signal (Methods, Supplementary Figure 9). Second, there was no association across the genome between S-pTDT ranking and either a) presence of an ASD-associated CNV (Supplementary Figure 10) or b) segmental duplication content of the region (Supplementary Figure 11). Third, we queried the specificity of the S-pTDT finding at 16p to ASD by performing an analogous analysis using a cohort of ADHD trios and an external ADHD GWAS; we did not replicate the finding in ADHD (Supplementary Figure 12). Finally, we do not see evidence that the ASD S-pTDT signal at 16p extends to the q-arm of chromosome 16 (Supplementary Figure 13). In summary, overtransmission of ASD polygenic risk at 16p is not driven by CNV carriers in our data, genomic structural features genome-wide, or as a cross-trait finding.

Finally, we analyzed the gene composition of 16p in relation to the 73 other 33Mb control regions and asked whether gene density could explain the S-pTDT signal (Methods). With 433 genes, 16p is the 3rd most gene dense region (**Figure 1F**). Furthermore, with 62 genes specifically expressed in the brain, 16p ranks 2nd highest relative to the other 33Mb control regions and has 37% more than predicted by the number of total genes --the greatest excess of any region (P = 0.001, **Figure 1F**). In contrast, 16p does not have a significant excess of genes implicated in ASD from exome associations studies (P = 0.44, Supplementary Figure 14).^31^ Given that 16p exhibits polygenic overtransmission and is gene dense, we tested the hypothesis that polygenic overtransmission reflects gene density. Across all 74 33Mb partitions, S-pTDT was not related to density of all genes (R = 0.025, P = 0.83), brain-specific genes (R = 0.084, P = 0.48), constrained genes (R = -0.023, P = 0.85), or genes associated with ASD via exome sequencing (R = 0.15, P = 0.21) (Supplementary Figure 15). Moreover, we do not observe a trend of gene dense regions having higher S-pTDT z-scores; for example, of the 13/74 partitions with more than 300 genes, the largest is 16p at z = 3.37 and the second largest is z = 1.32, much lower than 16p. Finally, we did not observe a relationship between the S-pTDT signal and density of fetal brain enhancers (R = 0.02, P = 0.88). This analysis suggests that while 16p is gene dense and enriched in brain-specific genes, these findings alone cannot explain a region’s degree of polygenic overtransmission.

### *In vitro* deletion of 16p11.2 causes decreased mean expression of genes on 16p

Whereas the 16p11.2 canonical CNV is only 0.7Mb in size, we observed S-pTDT signal across the entire p-arm of chromosome 16p. We hypothesized that the 16p11.2 deletion might have distal effects on gene expression across 16p. A previous report with endogenous (non-engineered) 16p11.2 deletion lines noted differential expression effects extending up to 5Mb from the 16p11.2 CNV.^19^ We sought to extend the analysis to the entire p-arm using engineered 16p11.2 deletions on an isogenic cellular background.

We used CRISPR/Cas9 to perform heterozygous deletion of the 16p11.2 CNV in iPSCs (n = 7 biological replicates), differentiated the cells into NGN2-induced neurons, and performed RNA-sequencing and differential expression analysis on the deletion lines relative to control neuronal lines without the 16p11.2 deletion (n = 6 biological replicates) (**Figure 2A**, Methods).^32^ We then asked whether, on average, the 200 neuronally expressed genes on 16p were differentially expressed in response to the 16p11.2 deletion (Methods). Genes on 16p had significantly lower expression in the deletion lines (n = 200 genes, mean log_2_(fold-change): -0.015, P = 0.02; mean fold-change t-statistic: -0.16, P = 0.01, **Figure 2B**); this observation remained significant after excluding the single gene (*TVP23A*) with decreased expression after Bonferroni correction. The deletion’s effect on 16p genes was different from the effect on all other 8,533 neuronally expressed genes in the genome (P = 0.02), whose expression was not on average changed by the deletion (P = 0.43) (**Figure 2C**). This analysis suggests that one of the most common ASD-associated CNVs is associated with transcriptional perturbation of genes in the surrounding region.

**Figure 2:**
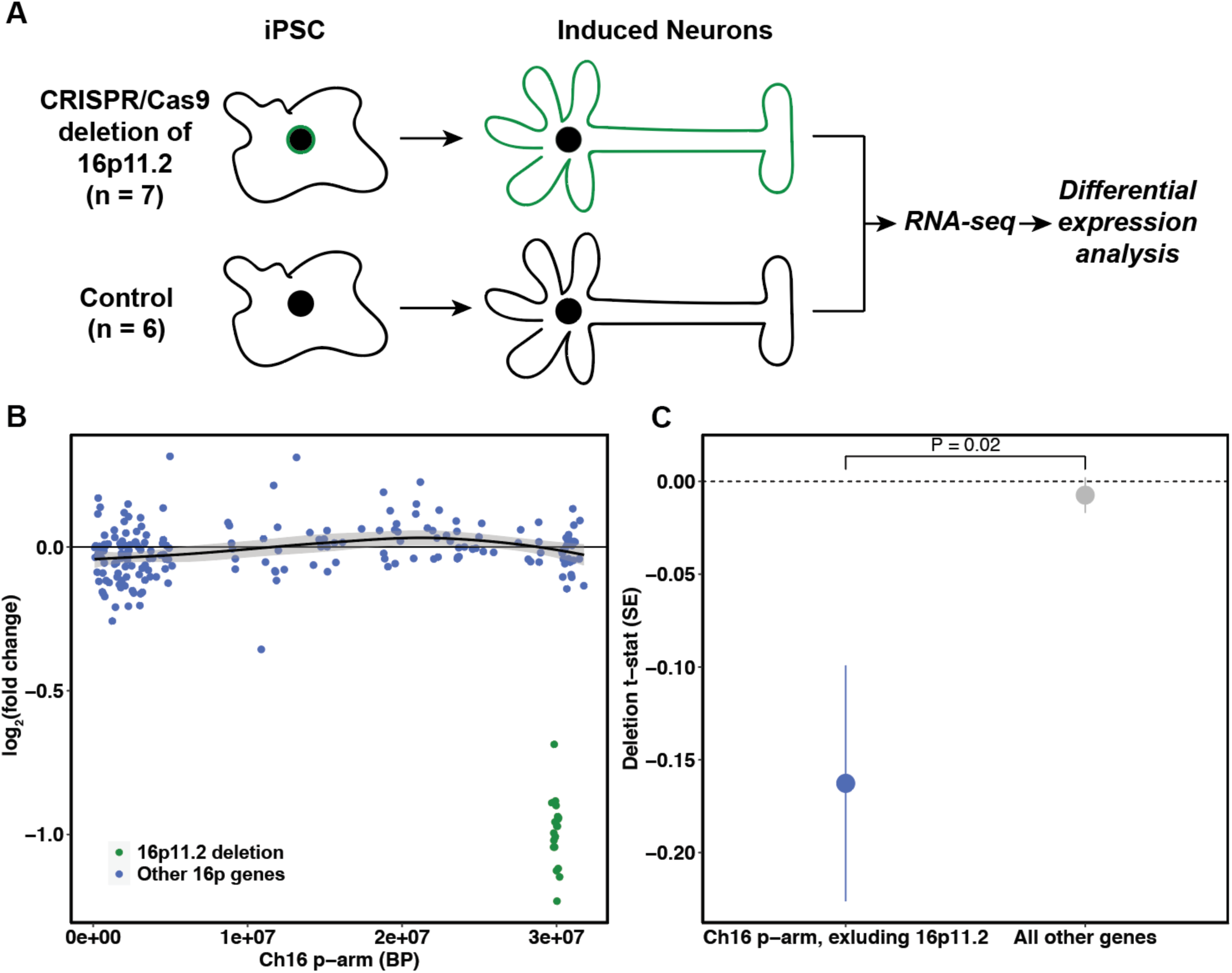
*in vitro* deletion of 16p11.2 causes decreased expression of 16p genes. **(A)** Experimental design of 16p11.2 *in vitro* deletion. Induced pluripotent stem cells undergo CRISPR-Cas9-mediated deletion of the 16p11.2 CNV region, differentiation into induced neurons and transcriptome profiling with RNA-seq (n = 7 biological replicates). Differential expression analysis compares these samples to controls (n = 6 biological replicates) without deletion of the CNV. **(B)** Differential expression of 16p brain genes after deletion of 16p11.2 CNV. Brain genes are defined based on above median normalized expression level of genes over all samples in analysis. Genes in the deletion region +/-0.1Mb are green, while all other genes on 16p are in blue. The y-axis is the log_2_ fold-change per gene. The trend line is for genes outside of the CNV (blue dots) with a 95% CI shaded in gray. **(C)** 16p11.2 CNV deletion causes decreased expression of brain genes on 16p but not of all other brain genes, where brain genes defined as in (B). Point estimates are of mean differential expression t-statistic for the group of genes +/-SE. P-value is from two-sample t-test comparing groups.

Recurrent deletions at 15q13.3 are also observed in ASD.^13,33–35^ To explore the specificity of our findings at 16p11.2, we explored the consequences of deletion of 15q13.3 in the same isogenic neuronal model (n = 11 heterozygous deletion replicates, n = 6 controls). In contrast to our 16p11.2 results, 15q13.3 was not associated with transcriptional perturbation of 100 neuronally expressed genes in the surrounding region (P = 0.42) and was not different than the effect on all other 8,087 neuronally expressed genes (P = 0.4) (Supplementary Figure 16, Methods). These results suggest that the transcriptional observations at 16p11.2 are not an artifact of the CRISPR/Cas9 construct, since the 15q13.3 and 16p11.2 models share CRISPR/Cas9 experimental procedure. These results also suggest that the regional transcriptional effects observed at 16p11.2 are not shared across all ASD-associated CNVs.

### 16p ASD PRS is associated with an average decrease in gene expression across 16p

Our analysis of 16p11.2 deletion lines suggests that this ASD rare variant risk factor causes transcriptional perturbation across 16p. Given that our S-pTDT analysis identified excess polygenic risk for ASD across the p-arm of chromosome 16, we tested the hypothesis that this common variant risk factor would also associate with decreased mean expression across 16p.

We analyzed paired genotype and expression data from three sources. First, we drew upon data from ongoing single-nucleus RNA-seq experiments on prefrontal cortex (BA46) from 122 European ancestry donors from the Harvard Brain Tissue Resource Center / NIH NeuroBioBank (HBTRC) (Supplementary Figure 17); we performed our primary analyses in the most abundant cell type (glutamatergic neurons) to maximize statistical power. Next, we analyzed paired genotype and bulk cortical RNA-sequencing from the CommonMind Consortium, split into two ancestry-specific subgroups (n = 194 individuals of African ancestry, n = 238 individuals of European ancestry) (Supplementary Figure 18).^36^ Both the HBTRC and CommonMind cohorts included donors with and without schizophrenia, and we controlled for case status in our analyses. Within each cohort, we constructed regional polygenic risk scores for ASD within the 33Mb partitions described above and regressed average regional gene expression on the regional polygenic risk score (Methods). To increase power, and to be consistent across data sets, we restricted each of the three analyses to the half of genes most strongly expressed in glutamatergic neurons in the HBTRC data (n = 8,878 genes).

Increased ASD PRS within 16p was associated with decreased expression in glutamatergic neurons of genes through the 16p region (**Figure 3A**; combined cohort per-gene permutation P-value: 0.048, Methods). Next, we compared the association between ASD PRS and mean gene expression observed at 16p to the strength of the same association in other 33Mb regions of the genome. Relative to the 73 other control regions, 16p had the second most negative association between regional PRS and mean gene expression (**Figure 3B**; beta = -0.025). Furthermore, 16p had by far the most consistently negative association between PRS and gene expression across the three cohorts (**Figure 3C**). In summary, we observe across independent cohorts that increased 16p ASD PRS is associated with an average decrease in gene expression within the partition.

**Figure 3:**
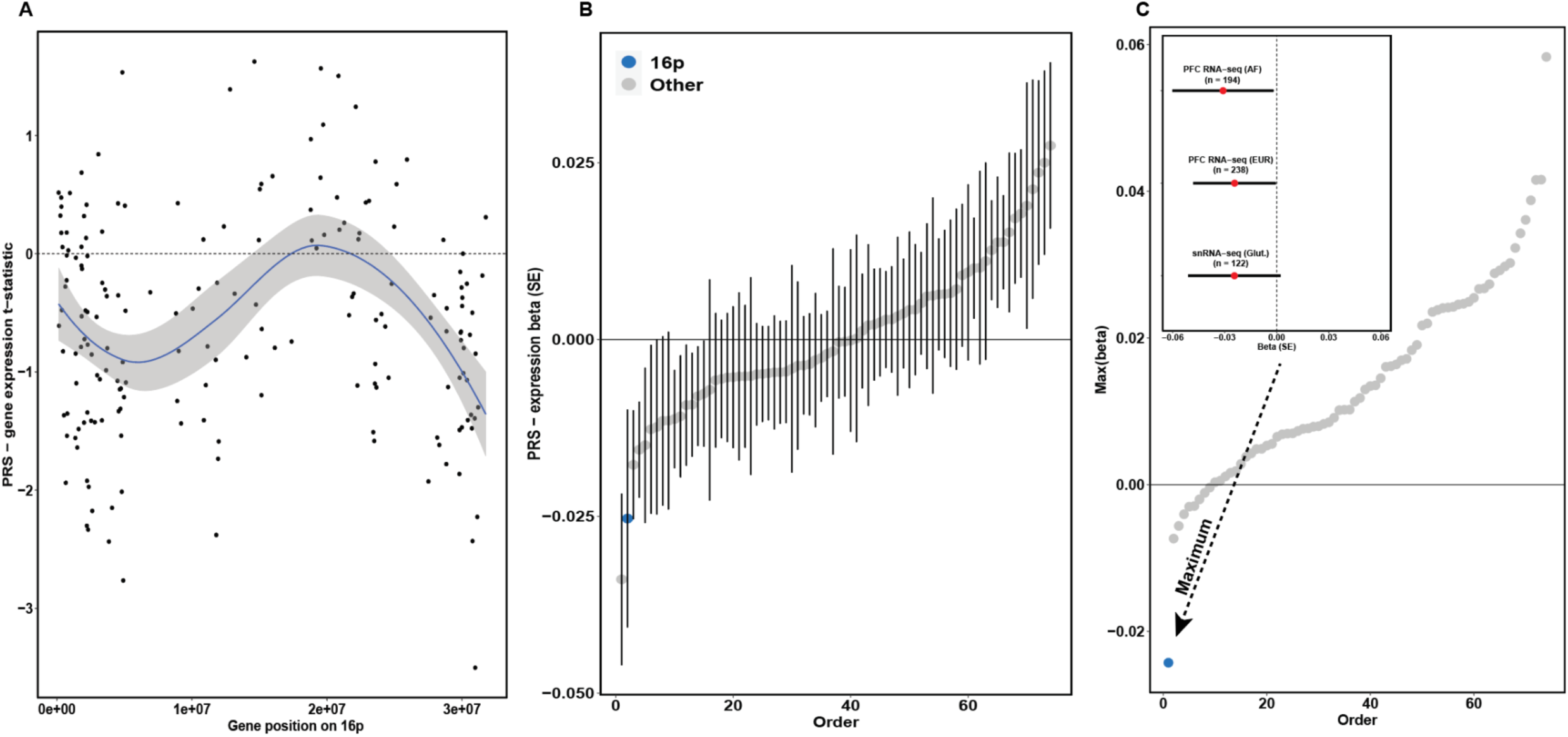
16p PRS is associated with decreased average expression of brain expressed genes on 16p. **(A)** Per gene association to 16p ASD PRS in combined analysis of HBTRC and CommonMind resources (n = 554 samples). Each point is a gene expressed in glutamatergic neurons (n = 183, Methods), the x-position its midpoint and the y-position its t-statistic from a linear model of normalized expression on 16p ASD PRS, controlling for donor schizophrenia case/control status, European ancestry, and single-cell/bulk expression measurement. The blue line is the mean trend across positions with a 95% confidence interval shaded in gray. **(B)** Association between average regional gene expression and regional PRS across 16p (blue point) and 73 other 33Mb control regions (gray points). Point estimates and error bars (SE) from regression of average regional gene expression on regional ASD PRS, controlling for donor schizophrenia case/control status, European ancestry, and single-cell/bulk expression measurement. **(C)** The 16p region exhibits the most consistent negative association between PRS and gene expression across the three cohorts compared to other 33Mb regions. Each point is a 33Mb region, with 16p marked in blue. The y-axis is the largest regression coefficient in the model described in (B), run individually for each of the three cohorts (HBTRC glutamatergic neurons n = 122 European ancestry samples; CommonMind (bulk cortical) n = 238 European ancestry samples; CommonMind (bulk cortical) n = 194 African ancestry samples). Inset: Association between mean gene expression and ASD 16p at 16p across the three cohorts.

### Chromatin contact at 16p associates with convergence of common and rare variant effects on gene expression

Given that both the 16p11.2 deletion and 16p ASD PRS are associated with decreased average gene expression in 16p, we asked whether these effects converged at the level of individual genes. We found a positive association between the per gene expression effects of the 16p11.2 deletion and the 16p ASD PRS across 168 glutamatergically expressed genes shared across both datasets (R = 0.19, P = 0.02, **Figure 4A**, Supplementary Figure 19). This observation suggests that the common variant 16p ASD PRS and the rare variant 16p11.2 deletion share downstream functional impact on gene expression. We also note that genes whose expression is decreased in response to both the 16p ASD PRS and the 16p11.2 deletion are enriched at the end of 16p (Ch 16: 0-5.2Mb, from here “telomeric region”, chi-squared P = 0.003 for negative t-statistic in both cohorts and telomeric region location, Methods). This telomeric region of chromosome 16 is extremely gene dense – with 182 genes, the 2nd most gene dense of 526 5.2Mb regions in the genome. As with 16p more broadly, it is enriched in genes specifically expressed in adult cortex (n = 33, 83% more than expected by chance, P = 2e-4).

**Figure 4:**
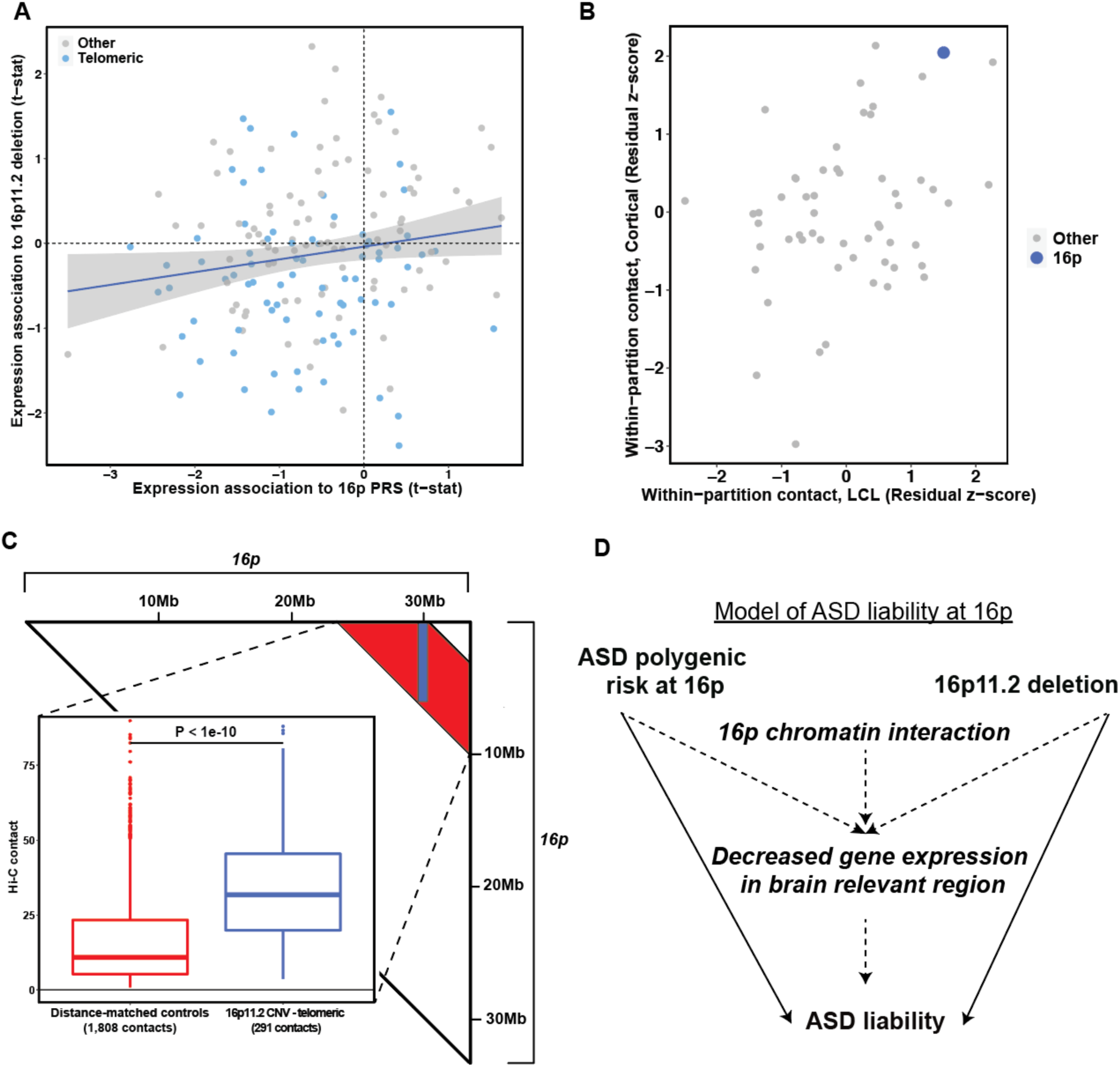
Integrative model of ASD liability at 16p. **(A)** The positive correlation of 16p genes in their association to 16p PRS and to the 16p11.2 *in vitro* deletion. The x-axis shows the association t-statistics from the all sample meta-analysis of 16p PRS and 16p gene expression. The y-axis shows the association t-statistics from the 16p11.2 *in vitro* deletion analysis. The shaded region is 95% CI. Genes are colored by their location on 16p; telomeric is defined as gene mid-point < 5.2Mb, while centromeric is all other genes on 16p. A single outlying point has been truncated from the plot for visualization; the untruncated plot is Supplementary Figure 19. **(B)** Hi-C analysis reveals elevated within-region chromatin interaction at 16p. Each point represents a 33Mb partition, with 16p colored blue. Both axes are in units of residual z-score, where the residual is from a linear model regressing out segmental duplication content and gene density from the mean within-region Hi-C contact value (Methods). The x-axis is a dataset of lymphoblastoid cell lines, while the y-axis is a dataset of mid-gestational cortical plate. **(C)** Hi-C analysis reveals elevated contact between 16p11.2 CNV and the 5.2Mb gene-dense telomeric region of 16p in mid-gestational cortical plate neurons. The triangle depicts the 16p contact matrix: the blue shaded region denotes contacts between the 16p11.2 CNV (29.5Mb-30.2Mb) and 0-5.2Mb telomeric region (n = 291 100kb x 100kb contacts), while the red shaded region are distance matched controls (n = 1,808 100kb x 100kb contacts). The inset shows the distribution of contact values for 16p11.2-telomeric vs. controls. The p-value is from a two-sample t-test comparing the distributions. **(D)** A model of ASD liability at 16p. Two independent genetic risk factors for ASD – the 16p11.2 deletion and polygenic variation at 16p – are located in a region of elevated 16p chromatin interaction and enriched in brain-specific expression, and are associated with coordinated decreased gene expression at 16p.

The correlation in transcriptional effects associated with the 16p PRS and the 16p11.2 deletion motivated us to explore genomic structural factors that could help to explain these coordinated effects across a large segment of the genome. We hypothesized that the 16p region may have increased within-region chromatin contact, which could explain the apparent non-independence of genetic and expression variation at megabase scale. To examine chromatin contact within 16p, we used two published Hi-C datasets: a dataset of lymphoblastoid cell lines (1Mb bin resolution)^37^ and a dataset from the primarily neuronal mid-gestational cortical plate (0.1 Mb bin resolution)^38^. Since our hypothesis pertained to average contact behavior over large regions of the genome – as opposed to more fine-grained analysis of topologically associated domains or gene-enhancer interactions – we analyzed the largest window available within each cohort to increase signal:noise ratio.^39^

The *i,j*^*th*^ entry of a Hi-C contact matrix estimates the degree of physical interaction between the *i*^*th*^ and *j*^*th*^ regions of genome. We estimated within 33Mb-partition contact as the mean of the off-diagonal values of the contact matrix. As segmental duplication content and gene density of the partition are associated with mean Hi-C estimates (Supplementary Figure 20), we regressed them out to yield a per-partition z-score which we interpret as the Hi-C regional contact corrected for these genomic features. We first noted that within region contact estimates are correlated across the two datasets (R = 0.31, P = 0.02, **Figure 4B**). The 16p partition exhibits high levels of within-region contact in both cohorts: the 4/74 highest partition in LCL lines (z-score: 1.50, 1.0Mb Hi-C resolution) and the 2/74 highest in the cortical plate dataset (z-score: 2.05, 0.1Mb Hi-C resolution). We hypothesize that this diffusely elevated within-region contact at 16p could facilitate the influence of regional polygenic effects on gene expression across 16p, via complex distal regulatory interactions.

Our analysis of the *in vitro* 16p11.2 deletion neurons (Figure 2B) revealed an effect of decreased gene expression at the gene-dense telomeric region of chromosome 16. We hypothesized that this is because the 0.7Mb 16p11.2 CNV has increased physical interaction with this telomeric region. Indeed, in mid-gestational cortical plate Hi-C data, the CNV-telomeric contacts (n = 291 100kb x 100kb contacts) are 2.9x more frequent than contacts between distance-matched control regions on 16p (n = 1,808 100kb x 100kb contacts, P < 1e-10, **Figure 4C**, Methods). In conclusion, these results suggest that the 3D conformation of 16p may mediate convergent ASD-related genetic effects on gene expression via regulatory interactions across megabases of separation.

## Discussion

Our observations motivate us to hypothesize the following model for ASD liability at 16p (**Figure 4D**): genetic liability for ASD emerges from the well-established 16p11.2 deletion, as well as an excess of common polygenic risk that is diffusely distributed across the region. Both of these risk factors are associated with a mean decrease in brain gene expression across 16p; their expression effects are correlated at the per gene level. We hypothesize that these transcriptional changes create ASD risk subsequent to the enrichment of brain specific genes in the region. We also hypothesize that the region’s elevated internal chromatin contact may facilitate the transcriptional convergence of these two distinct risk factors: a 0.7 Mb CNV and a 33Mb partition of common polygenic risk. This hypothesis is consistent with work demonstrating both single nucleotide^27^ and structural^24–26^ variation can cause transcriptional and chromatin perturbation. The distributed effect is also consistent with the results of a recent large-scale exome-sequencing study of ASD, which found that no single gene within the 16p11.2 locus was strongly associated with ASD.^8^ Our model adds to a literature of multi-gene^21^ and genetic network effects^19,22,23^ associated with the 16p11.2 CNV, and is the first to integrate common variation and chromatin architecture with 16p11.2 and the broader 33Mb 16p region.

Our analysis of large (e.g., 33Mb) regions of genome is non-canonical in complex trait genetics, contrasting with a common approach focused on mapping disease-associated variants to the genes through which they act.^4–6^ Existing approaches such as TWAS aggregate individually modest genetic effects on expression to associate genes with phenotype.^40^ Here, we both aggregate genetic effects on expression and aggregate effects across many genes in a region, further increasing power to observe modest effects. Regional analysis also allows new perspectives into gene function, including the observation of a region enriched in genes specifically expressed in brain, or enriched in within-region Hi-C contact. Our results suggest that chromatin landscapes can facilitate convergent genetic and transcriptional effects within large regions of the genome. This insight supports the viability of a new approach for extracting biological insight from genetic association data across large genomic regions.

Our observations raise many questions for future study. Why are the genetic and transcriptional associations at 16p related to ASD? On the one hand, we found that the region harbors an unusual concentration of genes specifically expressed in brain, but on the other, not an unusual number of genes implicated in ASD from exome association studies. We did not find a 16p signal in ADHD trios using the S-pTDT analysis, arguing against viewing 16p as equally relevant across neurodevelopmental traits. It is also possible that the genic relevance of the region will only become apparent through analysis of the biological networks into which 16p proteins interact and integrate; growing resources of protein-protein interaction data will facilitate this line of inquiry.^41^ The mean expression effects are modest, especially compared to the decrease in gene expression associated with heterozygous gene deletion such as that seen with the 16p11.2 CNV. Future studies will probe the biological consequence of modest expression change spread across many genes. This analysis also raises the question whether there are other regions of the genome where common and rare variation converge in a similar fashion with relevance either for ASD or for other traits. In conclusion, our analysis presents a novel statistical approach for partitioned polygenic association and uncovers surprising functional convergence of common and rare variant risk for ASD at 16p.

## Methods

### Generation of polygenic scores

For ASD PRS analysis in ASD trios, we used a GWAS from the iPSYCH collection in Denmark because there is no sample overlap with the ASD trio samples (19,870 cases, 39,078 controls) (Supplementary Table 1). For all other ASD PRS analysis (i.e. PRS - expression analyses), we used a meta-analysis of the same iPSYCH ASD samples, plus ASD samples from the Psychiatric Genomics Consortium (combined: 26,067 cases, 46,455 controls). For analysis of ADHD from the PGC, we used a non-overlapping iPSYCH-only ADHD GWAS (25,895 cases, 37,148 controls).

To generate polygenic scores weights, we first applied LDpred v1.11 on the marginal effect sizes from GWAS of our traits.^42^ We used LDpred under the infinitesimal genetic architecture model with LD reference from Hapmap 3 SNPs and an LD radius of 384 SNPs. All polygenic scores were calculated using the linear --score function in PLINK.^43^ Since LDpred estimates posterior causal effect sizes from GWAS marginal effect sizes, we include all SNPs in PRS analysis, including when constructing stratified polygenic scores for S-pTDT.

### ASD family cohorts

The collection and imputation of the Simons Simplex Collection (SSC) and Simons Foundation Powering Autism Research (SFARI) have been described previously (Supplementary Table 2).^12,44^ The ASD trios from the Psychiatric Genomics Consortium Autism group (PGC) are as described previously,^12^ with the modification of the inclusion of probands from multiplex families. We defined a European ancestry subset of PGC for analysis by generating principal components of ancestry using PLINK and by visual inspection relative to Hapmap reference populations (Supplementary Figure 1). We defined a family as European ancestry if both parents and proband were European ancestry by PC analysis (5,283 of 4,335 trios, 82%).

### Genome-wide pTDT

We performed genome-wide pTDT to assess power for stratified pTDT analyses in SSC, SPARK and PGC. We estimated polygenic transmission as described previously,^12^ with the exception of an adapted approach for the case/pseudocontrol genotypes in the PGC (Supplementary Note 1). Results for each of the three cohorts are listed in Supplementary Figure 2.

### Stratified pTDT (S-pTDT)

Stratified pTDT (S-pTDT) is identical to pTDT, except instead of testing for transmission of a polygenic score constructed from all SNPs, it tests for transmission of a polygenic score constructed from a subset of SNPs

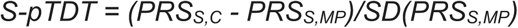

Where PRS_S,C_ is the stratified PRS of child C and PRS_S,MP_ is the mid-parent stratified PRS (average of the 2 parents). S-pTDT is a 1-sample t-test for whether the S-pTDT distribution has a mean different from 0. S-pTDT estimates for a given PRS for a given cohort is equal to the average S-pTDT value for all families in the cohort.

We created stratified polygenic scores by dividing SNPs into sets of equal sizes. We varied this partitioning in two ways to ensure an unbiased survey of regional transmission: first, we divided the SNPs into partitions of varying sizes (2,000 SNPs, 3,000 SNPs, 4,000 SNPs, 5,000 SNPs, 6,000 SNPs), and second, we started the partitioning either from the beginning or the end of the chromosome. This yielded 2,006 (often-overlapping) partitions (1,003 from the start of chromosomes, 1,003 from the end of chromosomes). Before partitioning we subsetted the PRS SNPs to those present in all three ASD trios cohorts (SSC, SPARK and PGC) to avoid bias from SNP missingness across partitions. We then estimated stratified PRS for each partition in each of the three cohorts using linear scoring (--score) in PLINK 1.9, and performed S-pTDT on each partition as described above.

Partition length and SNP count were predictive of overtransmission (Supplementary Figure 4). We regressed out expected overtransmission using a linear model: S-pTDT ∼ number_snps + partition_bp_length, and normalized the model residuals by the standard deviation of the model residual distribution. This procedure yields for each partition a residual z-score, which estimates the number of standard deviations the partition is over-or under-transmitted relative to expectation. If partitions included a gap between adjacent SNPs of greater than 1Mb, we adjusted the contribution of that gap down to 1Mb, which accounts for decay in LD but avoids inappropriately correcting the S-pTDT signal in the overtransmission model noted above. For analysis of 33Mb partitions, the S-pTDT z-score regressed out only SNP number, since basepair length of all partitions was the same.

### Supplemental analyses for 16p S-pTDT association

First, we confirmed that ASD-associated loci through GWAS were enriched in the S-pTDT distribution. We defined an ASD-associated locus as the five loci from the most recently published ASD GWAS reaching genome-wide significance from analysis of ASD alone.^11^

Two independent genetic risk factors for ASD – the 16p11.2 deletion and polygenic variation at 16p – are located in a region of elevated 16p chromatin interaction and enriched in brain-specific expression, and are associated with coordinated decreased gene expression at 16p.

Next, we analyzed the distribution of ASD-associated CNVs in the S-pTDT distribution. We identified ASD-associated CNVs from the set on SFARI Gene (https://gene.sfari.org/database/cnv/) and then identified the S-pTDT partitions with one of these CNVs within the boundary (Supplementary Figure 10). We also estimated the association between segmental duplication content and S-pTDT for the 33Mb partitions: we annotated each partition for segmental duplication rate by calculating the fraction of nucleotides in each partition that overlapped at least one segmental duplication per the UCSC Genome Browser.^45^ Coverage calculations were performed using BEDTools v2.30.0 (Supplementary Figure 11).^46^

To rule out the contribution of 16p CNV carriers to the S-pTDT signal, we repeated S-pTDT analysis in SSC + SPARK after removing trios where the proband carried an inherited or *de novo* neurodevelopmental - disorder associated CNV at 16p (we could not perform this analysis in PGC because we do not have exome sequencing for this cohort). We adopted a literature-based definition of neurodevelopmental -disorder associated from a recent ASD sequencing manuscript.^8^ Of the 5,048 trios in the SSC + SPARK analysis, we removed 51 (1.0%) with a qualifying CNV and repeated S-pTDT analysis (Supplementary Figure 9).

To evaluate the specificity of the S-pTDT finding on 16p, we performed an analogous analysis in ADHD. We used 1,634 European ancestry ADHD trios from the PGC and an external ADHD GWAS from the Danish iPSYCH collection with 25,895 cases and 37,148 controls.^47^ We partitioned the genome into blocks of 2,000, 3,000, 4,000, 5,000 and 6,000 SNPs as described above starting from the beginning of chromosomes and estimated S-pTDT for each partition. We then estimated a residual z-score, regressing out the number of SNPs and partition size as in the ASD analysis (Supplementary Figure 12).

We next evaluated whether the polygenic signal at 16p could be explained by a specific locus within the region. To perform this analysis, we partitioned 16p into approximately 1.5Mb LD-independent blocks as previously defined.^30^ We then estimated S-pTDT using the iPSYCH-only ASD summary statistics in SSC and SPARK for each of these 25 blocks (Supplementary Figure 8). To evaluate the contribution of individual loci, we estimated the decay in 16p S-pTDT signal as a function of removing the most over-transmitted remaining S-pTDT blocks. Specifically, we 1) estimated per block transmission 2) ranked the blocks from most to least overtransmission 3) estimated overtransmission using SNPs from all blocks 4) estimated overtransmission using SNPs from all blocks minus SNPs from the most associated remaining block 5) repeated Step 4 until only a single block remained (Figure 1E). For example, the first block (“Number of 16p Partitions Removed from PRS = 0”) includes all the 7,658 SNPs in the 16p PRS. The next block (“Number of 16p Partitions Removed from PRS = 1”) subtracts 287 SNPs from the most associated block in 16p, leaving this new block with 7,371 SNPs.

We next evaluated the regional polygenic signal of 16p relative to equally-sized comparison partitions across the genome. Since 16p spans approximately 33Mb of the genome, we constructed control partitions of 33Mb by starting at the beginning of chromosomes and defining adjacent 33Mb blocks. We defined the start coordinate of a chromosome by the minimum of (a) first SNP in 1000 Genomes Phase 3 EUR and (b) start position of first gene in gnomAD.^48,49^ Similarly, we defined the end coordinate of a chromosome by the maximum of (a) the last SNP in 1000 Genomes Phase 3 EUR and (b) and end position of the last gene in gnomAD. This approach yielded 74 partitions, including 16p. We performed S-pTDT using these boundaries by constructing stratified PRS from all SNPs within a given partition.

### Gene density

We first compiled a consensus gene list for gene density analyses. We defined this consensus list as the intersection of (a) autosomal genes with unique gene names and non-missing pLI constraint estimates from gnomAD and (b) genes with estimated specific expression in GTEx cortex (“Brain_Cortex”).^50^ This consensus list included 17,909 genes. We further annotated this list with the 102 genes implicated in ASD via exome-sequencing in a recent analysis.^31^ We then mapped genes to the above defined 33Mb boundaries if their gene body midpoint was located within the boundary. We built linear models predicting specific expression in cortex (top 10% of specific expression t-statistic) from density of all genes and calculated two-sided model p-values from the residual z-scores of the regression.

### CRISPR/Cas9 genome editing, cell model development, and differential expression analysis

#### Guide RNA design, iPSC culture and DNA transfection

16p11.2 CRISPR-engineered, isogenic 16p11.2 human induced pluripotent stem cell (hiPSC) lines with deletion of the 16p11.2 region were generated using the SCORE approach.^32^ CNV deletion boundaries were defined as Ch16 29,487,574-30,226,919 (GRCh37). Following single-cell isolation and screening, we retained CRISPR-treated lines harboring 16p11.2 copy number variants (CNVs) (n = 7) and two types of control lines including those were not exposed to CRISPR (n = 3) and those from CRISPR treatment without guide RNAs (n = 3). Briefly, for design of the optimal guide RNA, we first identified all possible 18–25mer guides with Jellyfish and performed a degenerate BLAST search to identify sequences that would uniquely target the 16p11.2 SDs, respectively, with no predicted off-target effects. The gRNA was cloned into pSpCas9(BB)-2A-Puro plasmid with a puromycin resistance marker (pX459, Addgene plasmid 48139) using a BbsI restriction site. Validation of the guide sequence in the gRNA vector was confirmed by Sanger Sequencing. Before transfection, all plasmids were purified from EndoFree Plasmid Maxi Kit according to the manufacturer’s instruction (Qiagen). Lines selected for differentiation underwent magnetic activated cell sorting (MACS) for expression of the TRA-1-60 cell surface marker for selection of pluripotent cells. Cells were separated using a MiniMACS Separator (Miltenyi Biotec, 130-090-312) with Anti-TRA-1-60 microbeads (Miltenyi Biotec, 130-100-832) following manufacturer instructions (∼2×10^6^ cells per line). TRA-1-60 positive cells were plated with Y-27632 dihydrochloride (10 µM), expanded, and cryopreserved using mFreSR. Cells within three passages of TRA-1-60 selection were used for differentiation.

Design of the 15q13.3 CNV deletion lines followed as above, with the deletion defined with boundaries Ch15 30,787,764-32,804,328 (GRCh37). N = 11 heterozygous deletion lines were created, with n = 6 control exposed to CRISPR construct but not to gRNA

#### Differentiation of induced Neurons (iNs)

TRA-1-60 positive hiPSCs were plated as single cells at 80% confluence on a Matrigel-coated 6-well plate with Y-27632 dihydrochloride. Polybrene (hexadimethrine bromide; Sigma, 107689) was added at 8 mg/mL within three hours of re-plating. Cells were incubated with polybrene for 10-15 minutes prior to the addition of lentivirus. Lentiviral constructs for directed differentiation of hiPSCs into iNs were made as described previously and added to polybrene-treated hiPSCs.^51^ Cells were incubated with lentivirus for 24 hours, followed by a media change with regular E8. At least 48 hours following single-cell re-plating, transduced hiPSCs were cryopreserved and passaged for expansion.

Transduced hiPSCs were expanded onto matrigel-coated T-25 flasks. Once all lines in a batch reached 70-80% confluence, cells were re-plated as single cells onto a new T-25 flask with Neural Maintenance Media (NMM) supplemented with Y-27632 dihydrochloride and 2µg/mL Doxycycline (Clontech, NC0424034) to begin induction of TetO gene driving Ngn2 expression and Puromycin resistance (Day 0). The NMM we use in this study is adopted from reference^52^. Twenty-four hours after re-plating, media was changed to NMM supplemented with 2 mg/L Doxycycline (Millipore Sigma, D9891) and 1 µg/mL Puromycin (Sigma), to begin selection of Ngn2-expressing cells (Day 1). Fresh NMM with Doxycycline and Puromycin was added to cells to continue selection on Days 2 and 3. On Day 4, cells were detached using Accutase (ThermoFisher, A1110501) and re-plated onto Poly-L-Ornithine (10 µg/ml; Sigma-Aldrich, P4957) / Laminin (5 µg/ml; Sigma-Aldrich, L2020)-coated plates with NMM supplemented with 2 mg/L Doxycycline, 10 mg/L human BDNF (Pro-Spec, CYT-207), and 10 mg/L human NT-3 (PeproTech, 450-03). Cells were counted prior to re-plating using a Countess II Automated Cell Counter (Invitrogen, AMQAF1000) with 2.5×10^5^ cells plated per well of a 12-well plate.

Following re-plating, iNs were not exposed to air and required half-media changes every other day. On Day 6, fresh NMM with Doxycycline, human BDNF, human NT-3, and 2 g/L Cytosine β-D-arabinofuranoside (Sigma, C1768-100MG) to prevent glial growth. On Day 8, a half-media change with fresh NMM with Doxycycline, BDNF, and NT-3 was conducted. For subsequent media changes (Day 10+), NMM supplemented with only BDNF and NT-3 was added until cells reached Day 24 of differentiation, at which time cells were dissociated for RNA-seq.

#### RNAseq library preparation and quality control

All RNA samples were extracted with Trizol reagent according to the manufacturer’s instruction (Invitrogen). RNA sample quality (based on RNA Integrity Number, RIN) and quantity were determined on an Agilent 2200 TapeStation and between 500-100 ng of total RNA was used to prepare libraries. iNs RNASeq libraries were prepared with Illumina’s TruSeq Stranded mRNA Library Kit, which used polyA capture to enrich mRNA, followed by stranded reverse transcription and chemical shearing to make appropriate stranded cDNA inserts for the library. Libraries were finished by adding both sample-specific barcodes and adapters for Illumina sequencing followed by between 10-15 rounds of PCR amplification. Final concentration and size distribution of libraries were evaluated by 2200 TapeStation and/or qPCR, using Library Quantification Kit (KK4854, Kapa Biosystems), and multiplexed by pooling equimolar amounts of each library prior to sequencing. RNAseq libraries were sequenced on multiple lanes of an Illumina HiSeq 2500 platform.

Quality of sequence reads was assessed by fastQC (version 0.10.1).^53^ Gene-based counts were generated by aligning sequence reads to the human reference genome, GRCh37 (v75) and relying on the Ensembl gene annotations of these reference genomes using STAR (version 2.4.2a), with parameters “-- outSAMunmapped Within --outFilterMultimapNmax 1 --outFilterMismatchNoverLmax 0.1 -- alignIntronMin 21 --alignIntronMax 0 --alignEndsType Local --quantMode GeneCounts -- twopassMode Basic”.^54^ Quality of alignments was assessed by custom scripts utilizing Picard Tools (https://broadinstitute.github.io/picard/), RNASeQC^55^ and SamTools^56^. Further exploratory analyses including clustering and principal component analyses (PCA) were implemented in R (version 3.4) using DESeq2 (version 1.18.1)^57^ and custom scripts.

#### Differential gene expression analysis

Differential expression (DE) analyses were performed on genes that passed the expression threshold in a given comparison using R/Bioconductor packages DESeq2 (v. 1.18.1).^57^ To determine genes that passed the expression threshold for a particular comparison, count-per-million (cpm) expression values of genes across the samples used in the comparison were calculated. Cpm of i^th^ gene in sample j was defined as 10^6^ x C_i_ / LS_j_, where C_i_ is raw counts of i^th^ gene and LS_j_ is the library size of j^th^ sample. Total number of uniquely mapped reads reported by STAR aligner for a given sample was taken as the library size of that sample. Next, cpm expression threshold for a given comparison was calculated for 10 counts, using the equation 10^6^ x 10 / median(LS), where median(LS) is the median value of library sizes of samples used in the comparison. All the genes with expression values in cpm equal or greater than the cpm expression threshold in at least 50% of samples in either condition (e.g. deletion or wild-type) were further analyzed in the DE analysis by comparing CNV type deletion with the corresponding wild-type samples. To account for unknown sources of variation in the expression data, surrogate variables (SVs) were estimated for each comparison using R/Bioconductor package the Surrogate Variable Analysis (SVA version 3.26) by setting ∼ genotype as the full model and ∼ 1 as the reduced model.^58,59^ Differential expression t-statistics were defined as DE effect size / DE standard error.

### PRS - expression analyses

#### Harvard Brain Tissue Resource Center / NIH NeuroBioBank *(Single-nucleus RNA-seq)*

We generated paired genotype and single-nucleus expression profiles from dorsolateral prefrontal cortex from deceased donors from the Harvard Brain Tissue Resource Center / NIH NeuroBioBank. The generation of expression profiles will be described in detail in a forthcoming manuscript from Ling et al. In brief, we developed and optimized a workflow for creating and analyzing “brain nuclei villages” – nuclei sampled from brain tissue (dorsolateral prefrontal cortex, DLPFC, BA46) from 20 different donors per village. In this workflow, we start by dissecting a defined amount of tissue from each donor, obtaining a similar mass of tissue from each specimen while being careful to represent all cortical layers. The frozen tissue samples are then immediately pooled for simultaneous isolation of their nuclei; all subsequent processing steps – including nuclear isolation, encapsulation in droplets, and preparation of snRNA-seq libraries – involve all of the donors together. This “Dropulation” workflow allows us to minimize experimental variability, including any technical effects on mRNA ascertainment and any effects of cell-free ambient RNA. Each nucleus in these experiments was then re-assigned to its donor-of-origin using combinations of hundreds of transcribed SNPs; though the individual SNP alleles are shared among many donors, the combinations of many SNPs are unique to each donor in the cohort. Nuclei were assigned to 7 major cell classes (astrocytes, endothelial cells, GABAergic neurons, glutamatergic neurons, microglia, oligodendrocytes, polydendrocytes) by global clustering and identification of marker genes expressed in each cluster. Median cell type proportions were: glutamatergic neurons 47.9%, GABAergic neurons 18.8%, astrocytes 13.5%, oligodendrocytes 8.0%, polydendrocytes 5.2%, microglia 1.5%, endothelia 1.0%; we performed subsequent analyses using the most abundant cell type (glutamatergic neurons) to maximize power. The cell-type specific gene-by-donor expression matrices were processed with VST normalization.^60^

We performed a number of pre-association QC steps. The majority of genotyped samples were European ancestry (1707/1770, 96%), and we identified these samples for downstream analysis using PCA (Supplementary Figure 17). Next, we identified any samples as expression outliers with mean expression > 3 SD from the cohort mean (3/125 samples). This yielded a final EUR subset of 122 samples.

We then analyzed the relationship between mean gene expression and regional ASD PRS in these samples. Using the 33Mb partitions described above, we estimated a stratified ASD PRS in these samples using the largest ASD GWAS (iPSYCH + PGC, see Supplementary Table 1) using Plink --score and genotype QC (SNP missingness < 1%, MAF > 0.1%, imputation INFO > 95%). For gene expression profiles, we restricted analysis to the top 50% of genes by per sample normalized raw count expression, yielding 8,878 genes. Within each 33Mb partition, we performed per gene regional PRS to gene expression association with the linear model: gene VST expression ∼ regional PRS + schizophrenia case/control. Each regression produces a t-statistic for the regional PRS coefficient, which estimates the normalized association of the regional PRS with gene expression in cis. After performing the regression for each gene in the partition, we calculate the mean t-statistic for genes in the partition (Figure 3B, Supplementary Figure 19).

#### CommonMind (Bulk cortical RNA-seq)

We next analyzed paired genotype and bulk dorsolateral prefrontal cortex expression data from donors in the CommonMind consortium. Generation of expression count matrices is described in the CommonMind publication.^36^ Within CommonMind, we restricted analysis to donors from the NIMH Human Brain Collection Core (HBCC) and the University of Pittsburgh (PITT) biobanks due to previous analysis demonstrating increased consortance with the Dropulation data (Ling et al, forthcoming). We performed variance stabilization on the count matrices separately in HBCC and PITT using the SCTransform package in Seurat with the goal of closely paralleling the primary analysis in Dropulation (parameters: do.scale=FALSE, do.center=FALSE, return.only.var.genes = FALSE, seed.use = NULL, n_genes = NULL). The CommonMind collection is ancestrally heterogeneous; the two largest groups are African and European ancestry donors. Accordingly, we used PCA to identify donors of African ancestry (n = 194) and European ancestry (n = 238) and subsequently analyzed each separately (Supplementary Figure 18). For consistency with the Dropulation sample, we restricted analysis to donors diagnosed with schizophrenia or controls.

As in the Harvard Brain Tissue Resource, we estimated the association between regional polygenic scores and average shifts in regional gene expression. For each ancestral cohort, we associated ASD regional PRS to the normalized expression of genes in the region: expression ∼ regional PRS + schizophrenia case/control. For consistency, we restricted analysis to the same genes analyzed in the Harvard Brain Tissue Resource analysis described above.

#### Per cohort association and meta-analysis

We performed two classes of local PRS - gene expression association. The first class is per-gene association, as in Figure 3A. The second is average gene association, as in Figure 3B-C. To be consistent across data sets, we restricted all analyses to the half of genes most expressed in glutamatergic neurons in the HBTRC data (n = 8,878 genes). The association in Figure 3A is per-gene association meta-analyzed across the three cohorts. We combined individual level expression and genotype PRS across the three cohorts; before concatenating the polygenic scores and expression matrices used in the individual cohort analyses, we within-cohort scaled per gene gene expression and per partition PRS to mean = 0 and SD = 1. Per gene association followed the linear model: gene expression ∼ regional PRS + schizophrenia case/control + ancestry (binary for yes/no African ancestry) + single_cell (binary yes/no). The association t-statistic is from the regional PRS covariate. For maximum power to detect mean effects, we assessed significance of the mean PRS-expression association using permutation. Specifically, we calculated the mean(t-statistic) in 16p, then shuffled the PRS-donor IDs, performed association, then calculated the mean(t-statistic), repeated 5,000 times. The permutation p-value is the number of times the observed PRS was more negative than the permuted PRS. For the second class of association, we first averaged the gene expression per partition, then performed association. For per cohort association, the model is: mean expression of gene ∼ regional PRS + schizophrenia case/control. For combined analysis, the model is mean gene expression ∼ regional PRS + schizophrenia case/control + ancestry (binary for yes/no African ancestry) + single_cell (binary yes/no).

### Hi-C analysis

#### LCL lines

Per chromosome Hi-C count matrices were downloaded from http://hic.umassmed.edu/ at 1Mb resolution for GM06990 lymphoblastoid cell line.^37^ Since the count matrices were built in hg18, we converted the 33Mb partitions from hg19 to hg18 using the NCBI Genome Remapping Service (www.ncbi.nlm.nih.gov/genome/tools/remap). We matched the boundaries of the 33Mb partitions with their closest 1Mb cutoffs in the count matrix. For this analysis, we did not analyze partitions spanning centromeres, yielding 56 partitions for analysis. For each partition, we estimated raw within partition contact frequency as the mean of the off-diagonal elements of the Hi-C count matrix.

#### Mid-gestational cortical plate

We download 0.1Mb resolution Hi-C contact matrices from NCBI GEO from a resource of mid-gestational cortical plate samples from 3 donors.^38^ As above, we mapped the boundaries of the 33Mb partitions to the 0.1Mb boundaries of the Hi-C matrix. In contrast to the LCL lines, the diagonal elements were zeroed out; thus, the estimated raw within partition contact frequency was estimated as the mean of all elements of the matrix. We analyzed the same 56 partitions as in the LCL analysis.

#### Contact model

Raw within-partition contact frequency varies with gene density and segmental duplication contact (Supplementary Figure 21). In the LCL lines, this covariance is likely due to increased number of Hi-C reads mapping to regions with increased segmental duplication content. In cortical lines, there are large chunks of zeroed out elements of the contact matrix, rates of which correlate strongly with segmental duplication content, likely due to intentional zeroing of elements in regions that are difficult to map due to segmental duplication content. Gene density remains a significant predictor of contact frequency after conditioning on segmental duplication content, motivating us to condition on gene density as well and to extract normalized residuals from the following model: contact frequency ∼ gene density + segmental duplication content. Our primary analysis in Figure 4B reports these normalized residuals for each partition.

#### 16p11.2 CNV - telomeric region analysis

We next analyzed the chromatin contact between the 16p11.2 CNV and the distal gene-dense start of chromosome 16. We performed this analysis in the mid-gestational cortical plate data given its higher resolution of 100kb. We defined the telomeric region based on the gene dense segment at the start of chromosome 16, from 0Mb to the closest 100kb segment after the endpoint of the brain expressed gene in that window in the 16p11.2 deletion dataset (5.2 Mb). We defined the 16p11.2 CNV as Ch16: 29.5 - 30.2Mb. To define control contact regions, we first calculated the minimum (24.3Mb) and maximum (30.2Mb) distances spanned by the contact matrix defined by 0-5.2Mb (telomeric region) and 29.5-30.2Mb (16p11.2 CNV). We then defined control contacts on 16p as all contacts of distance greater than 24.3 or less than 30.2 that were not located in the telomeric-CNV contact matrix described above. These control regions have an intuitive geometric interpretation in the contact matrix as all of the contacts within the equidistant diagonal lines (forming a trapezoid, see Figure 4C). These results are robust to inclusion of elements of the contact matrix with “0”, which likely reflect segmental duplication rich regions (telomeric-CNV vs. control p-value < 1e-10 for both).

## Data Availability

Data is publicly available, or will be made available from principal investigators upon publication of the manuscript.

## Additional Information

## Acknowledgements

This work was generously supported by the Simons Foundation Autism Research Initiative (704413 to Elise Robinson and Luke O’Connor), the National Institute of Mental Health (F30MH129009 to Daniel Weiner, R01MH111813 to Elise Robinson, U01MH119689 to Elise Robinson, R01MH115957 to Michael Talkowski, 1R01MH124851-01 to Anders D. Børglum), the National Institute of General Medical Sciences (T32GM007753 to Ajay Nadig), the National Institute of Child Health and Development (R01HD096326 to Michael Talkowski), the National Institute of Neurological Disorders and Stroke (R01NS093200 to Michael Talkowski), the National Library of Medicine (T15LM007092 to Daniel Weiner), the Stanley Center for Psychiatric Research (to Steve McCarroll), the Lundbeck Foundation (R102-A9118, R155-2014-1724, and R248-2017-2003 to iPSYCH), the Novo Nordisk Foundation (to the Danish National Biobank) and the Universities and University Hospitals of Aarhus and Copenhagen. High-performance computer capacity for handling and statistical analysis of iPSYCH data on the GenomeDK HPC facility was provided by the Center for Genomics and Personalized Medicine and the Centre for Integrative Sequencing, iSEQ, Aarhus University, Denmark in a grant to Anders D. Børglum. Human tissue was obtained from the NIH NeuroBioBank.

We would like to thank all the families who participated in the cohorts included in this analysis, without whom this work would not be possible. We would also like to acknowledge Ryan Collins and Raymond Walters for their assistance with these analyses. Finally, we would like to thank the following members of the ADHD Working Group of the Psychiatric Genomics Consortium: A.R. Hammerschlag, Alejandro Corsico, Alexandra Havdahl, Alexandre Todorov, Alice Charach, Allison Ashley-Koch, Alysa Doyle, Amaia Hervas, Ana Miranda, Anders Borglum, Andre Scherag, Anita Thapar, Anna Rommel, Anna Starnawska, Anne Wheeler, Aribert Rothenberger, Aurina Arnatkeviciute, Barbara Franke, Ben Neale, Calwing Liao, Catharina Hartman, Christie Burton, Christine Cornforth, Cristina Sanchez, Danielle Posthuma, Felecia Cerrato, Fernando Mulas, Franziska Degenhardt, Glaucia Chiyoko Akutagava Martins, Gun Peggy Stromstad Knudsen, H.C. Steinhausen, Hakon Hakonarson, Hans-Christoph Steinhausen, Herber Roeyers, Hyo-Won Kim, Ian Gizer, Irwin Waldman, Isabell Brikell, Jennifer Crosbie, Jessica Agnew-Blais, Joanna Martin, Joel Gelernter, Johannes Hebebrand, Josep Antoni Ramos-Quiroga, Joseph Biederman, Joseph Sergeant, Juanita Gamble, Julia Pinsonneault, Jurgen Deckert, Kate Langley, Li Yang, Lindsey Kent, Luis Rohde, Manuel Mattheisen, Maria Jes’s Arranz Calderun, María Soler Artigas, Marta Ribases, Meg Mariano, Michael Gill, Mick O’Donovan, Miguel Casas, Monica Bayes, Nick Martin, Niels Peter Ole Mors, Nigel Williams, Nina Roth Mota, Ole Andreas Andreassen, Pak Sham, Patrick Sullivan, Paul Arnold, Paul Lichtenstein, Paula Rovira, Peter Holmans, Philip Asherson, Preben Bo Mortensen, Rachel Guerra, Raymond Walters, Richard Anney, Richard Ebstein, Richard Karlsson Linnér, Ridha Joober, Robert Oades, Russell Schachar, Sarojini M. Sengupta, Stefan Johansson, Stephanie H. Witt, Steve Nelson, Susan Smalley, Susann Scherag, Tetyana Zayats, Thomas Werge, Tim Silk, Tinca Polderman, Tony Altar, Veera Manikandan, Yanil Zhang, Yorgos Athanasiadis and Yufeng Wang. Support for title page creation and format was provided by AuthorArranger, a tool developed at the National Cancer Institute.

## Author contributions

D.J.W. conducted analyses. D.J.W, E.L., S.E., D.J.C.T., R.Y., J.G., J.F., C.C., N.B., S.B., D.M.H., J.B.G. and A.D.B generated data. D.J.W., E.L., S.E., D.J.C.T. and R.Y. designed experiment and tools. D.J.W., E.L., A.N., E.Z.M., J.S., L.J.O., A.D.B., M.E.T., S.A.M. and E.B.R. aided in interpretation of data. E.B.R. supervised the research. D.J.W. and E.B.R. wrote the manuscript.

### IRB

This study was reviewed and approved by Partners Human Research of Partners HealthCare. The study name is Molecular Study of Cognitive and Behavioral Variation (IRB: 2015P002376). The Principal Investigator is Elise Robinson. The iPSYCH study was approved by the Danish Data Protection Agency and the Scientific Ethics Committee in Denmark.

### Consortium authorship

#### iPSYCH Consortium

Preben B. Mortensen^1,2,3^, Thomas Werge^1,4,5^, Ditte Demontis^1,6,7^, Ole Mors^1,8^, Merete Nordentoft^1,9^, Thomas D. Als^1,6,7^, Marie Bækvad-Hansen^1,10^, Anders Rosengren^1,4^

^1^The Lundbeck Foundation Initiative for Integrative Psychiatric Research, iPSYCH, Aarhus, Denmark, ^2^National Centre for Register-Based Research, Aarhus University, Aarhus, Denmark, ^3^Centre for Integrated Register-based Research, Aarhus University, Aarhus, Denmark, ^4^Institute of Biological Psychiatry, MHC Sct. Hans, Mental Health Services Copenhagen, Roskilde, Denmark, ^5^Department of Clinical Medicine, University of Copenhagen, Copenhagen, Denmark, ^6^Department of Biomedicine (Human Genetics) and iSEQ Center, Aarhus University, Aarhus, Denmark, ^7^Center for Genomics and Personalized Medicine (CGPM), Aarhus University, Aarhus, Denmark, ^8^Psychosis Research Unit, Aarhus University Hospital, Aarhus, Denmark, ^9^Mental Health Services in the Capital Region of Denmark, University of Copenhagen, Copenhagen, Denmark, ^10^Center for Neonatal Screening, Department for Congenital Disorders, Statens Serum Institut, Copenhagen, Denmark

### ASD Working Group of the Psychiatric Genomics Consortium

Alexandra Havdahl^1^, Anders Rosengren^2,3^, Anders D. Børglum^4,5,2^, Anne Hedemand^5^, Aarno Palotie^6^, Aravinda Chakravarti^7^, Elise B. Robinson^8,9,10^, Dan Arking^11^, Arvis Sulovari^12^, Anna Starnawska^5^, Bhooma Thiruvahindrapuram^13^, Christiaan deLeeuw^14^, Caitlin Carey^8,9,15^, Christine Ladd-Acosta^16^, Celia van der Merwe^15^, Bernie Devlin^17^, Edwin H Cook^18^, Evan Eichler^19^, Elisabeth Corfield^20^, Gwen Dieleman^21^, Gerard Schellenberg^22^, Jakob Grove^4,5,23,2^, Hakon Hakonarson^24^, Hilary Coon^25^, Isabel Dziobek^26^, Jacob Vorstman^27^, Jessica Girault^28^, Jack M. Fu^8,29,30,9^, James S. Sutcliffe^31^, Jinjie Duan^5^, John Nurnberger^32^, Joachim Hallmayer^33^, Joseph Buxbaum^34^, Joseph Piven^28^, Laren Weiss^35^, Lea Davis^36^, Magdalena Janecka^37^, Manuel Mattheisen^2^, Matthew W. State^35^, Michael Gill^38^, Mark Daly^15^, Mohammed Uddin^39^, Ole Andreassen^40^, Peter Szatmari^27^, Phil Hyoun Lee^8^, Richard Anney^41^, Stephan Ripke^42^, Kyle Satterstrom^8^, Susan Santangelo^43^, Susan Kuo^8^, Ludger Tebartz van Elst^44^, Thomas Rolland^45^, Thomas Bougeron^45^, Tinca Polderman^46^, Tychele Turner^47^, Jack Underwood^48^, Veera Manikandan^49^, Vamsee Pillalamarri^11^, Varun Warrier^50^

^1^Department of Psychology, University of Oslo, Oslo, Norway, ^2^The Lundbeck Foundation Initiative for Integrative Psychiatric Research, iPSYCH, Aarhus, Denmark, ^3^Institute of Biological Psychiatry, MHC Sct. Hans, Mental Health Services Copenhagen, Roskilde, Denmark, ^4^Center for Genomics and Personalized Medicine (CGPM), Aarhus University, Aarhus, Denmark, ^5^Department of Biomedicine (Human Genetics) and iSEQ Center, Aarhus University, Aarhus, Denmark, ^6^Institue for Molecular Medicine, University of Helsinki, Helsinki, Finland, ^7^Center for Human Genetics and Genomics, New York University, New York, USA, ^8^Stanley Center for Psychiatric Research, Broad Institute of MIT and Harvard, Cambridge, MA, USA, ^9^Center for Genomic Medicine, Massachusetts General Hospital, Boston, MA, USA, ^10^Department of Psychiatry, Massachusetts General Hospital and Harvard Medical School, Boston, MA, USA, ^11^McKusick-Nathans Institute of Genetic Medicine, Johns Hopkins, Baltimore, MD, USA, ^12^Cajal Neuroscience, Seattle, WA, USA, ^13^The Centre for Applied Genomics, The Hospital for Sick Children, Toronto, Canada, ^14^Department of Complex Trait Genetics, VU University, Amsterdam, Netherlands, ^15^Analytic and Translational Genetics Unit, Massachusetts General Hospital, Boston, MA, USA, ^16^Bloomberg School of Public Health, Johns Hopkins, Baltimore, MD, USA, ^17^Department of Psychiatry, University of Pittsburgh, Pittsburgh, PA, USA, ^18^Department of Psychiatry, University of Illinois at Chicago, Chicago, IL, USA, ^19^Department of Genome Sciences, University of Washington, Seattle, WA, USA, ^20^Department of Mental Disorders, Norwegian Institute of Public Health, Oslo, Norway, ^21^Department of Child and Adolescent Psychiatry, Sophia Childrens Hospital, Rotterdam, Netherlands, ^22^Department of Pathology and Laboratory Medicine, University of Pennsylvania School of Medicine, Philadelphia, PA, USA, ^23^Bioinformatics Research Centre, Aarhus University, Aarhus, Denmark, ^24^The Center for Applied Genomics and Division of Human Genetics, University of Pennsylvania School of Medicine, Philadelphia, PA, USA, ^25^Department of Psychiatry, University of Utah, Salt Lake City, UT, USA, ^26^Department of Psychology, Humboldt Universitat Berlin, Berlin, Germany, ^27^Department of Psychiatry, University of Toronto, Toronto, Canada, ^28^Department of Psychiatry, UNC School of Medicine, Chapel Hill, NC, USA, ^29^Department of Neurology, Massachusetts General Hospital and Harvard Medical School, Boston, MA, USA, ^30^Program in Medical and Population Genetics, Broad Institute of MIT and Harvard, Cambridge, MA, USA, ^31^Center for Human Genetics Research, Vanderbilt University, Nashville, TN, USA, ^32^Department of Psychiatry, Indiana University School of Medicine, Indianapolis, IN, USA, ^33^Department of Psychiatry, Stanford University, Stanford, CA, USA, ^34^Department of Neuroscience, Icahn School of Medicine at Mount Sinai, New York, NY, USA, ^35^Department of Psychiatry, University of California, San Francisco, San Francisco, CA, USA, ^36^Vanderbilt Genetics Institute, Vanderbilt University Medical Center, Nashville, TN, USA, ^37^Department of Psychiatry, Icahn School of Medicine at Mount Sinai, New York, NY, USA, ^38^Department of Psychiatry, Trinity College Dublin, Dublin, Ireland, ^39^College of Medicine, Mohammed Bin Rashid University of Medicine and Health Sciences, Dubai, UAE, ^40^Institute of Clinical Medicine, University of Oslo, Oslo, Norway, ^41^Division of Psychological Medicine and Clinical Neurosciences, Cardiff University, Cardiff, UK, ^42^Department of Psychiatry and Psychotherapy, Charite, Berlin, Germany, ^43^Center for Psychiatric Research, Maine Medical Center Research Institute, Portland, ME, USA, ^44^Department of Psychiatry and Psychotherapy, University of Freiburg, Freiburg, USA, ^45^Departement of Neuroscience, Institut Pasteur, Paris, France, ^46^Department of Child and Adolescent Psychiatry, University of Amsterdam, Amsterdam, Netherlands, ^47^Department of Genetics, University of Washington School of Medicine in St. Louis, St. Louis, MO, USA, ^48^Cardiff University, Cardiff, UK, ^49^Regeneron, White Plains, NY, USA, ^50^Department of Psychiatry, University of Cambridge, Cambridge, UK

#### ADHD Working Group of the Psychiatric Genomics Consortium

Alexandra Philipsen^1^, Andreas Reif^2^, Anke Hinney^3^, Bru Cormand^4,5,6,7^, Claiton H.D. Bau^8,9^, Diego Luiz Rovaris^10^, Ditte Demontis^11,12,13^, Edmund Sonuga-Barke^14^, Elizabeth Corfield^15,16^, Eugenio Horacio Grevet^17,18^, Giovanni Salum^17,19^, Henrik Larsson^20,21^, Jan Buitelaar^22^, Jan Haavik^23,24^, James McGough^25^, Jonna Kuntsi^14^, Josephine Elia^26,27^, Klaus-Peter Lesch^28,29,30^, Marieke Klein^31^, Mark Bellgrove^32^, Martin Tesli^15^, Patrick WL Leung^33^, Pedro M. Pan^34^, Soren Dalsgaard^35,36,37^, Sandra Loo^25^, Sarah Medland^38^, Stephen V. Faraone^39^, Ted Reichborn-Kjennerud^40,41^, Tobias Banaschewski^42^, Ziarih Hawi^32^

^1^Department of Psychiatry and Psychotherapy, University Hospital Bonn, Bonn, Germany, ^2^Department for Psychiatry, Psychosomatic Medicine and Psychotherapy, University Hospital Frankfurt, Frankfurt, Germany, ^3^Department of Child and Adolescent Psychiatry, University Hospital Essen, Essen, Germany, ^4^Department of Genetics, Microbiology and Statistics, University of Barcelona, Barcelona, Catalonia, Spain, ^5^Centro de Investigación Biomédica en Red de Enfermedades Raras (CIBERER), Spain, ^6^Institut de Biomedicina de la Universitat de Barcelona (IBUB), Barcelona, Catalonia, Spain, ^7^Institut de Recerca Sant Joan de Déu (IR-SJD), Barcelona, Catalonia, Spain, ^8^Department of Genetics, Universidade Federal do Rio Grande do Sul, Porto Alegre, Brazil, ^9^ADHD and Developmental Psychiatry Programs, Hospital de Clínicas de Porto Alegre, Porto Alegre, Brazil, ^10^Instituto de Ciencias Biomedicas, Universidade de Sao Paulo, São Paulo, Brazil, ^11^The Lundbeck Foundation Initiative for Integrative Psychiatric Research, iPSYCH, Aarhus, Denmark, ^12^Department of Biomedicine (Human Genetics) and iSEQ Center, Aarhus University, Aarhus, Denmark, ^13^Center for Genomics and Personalized Medicine (CGPM), Aarhus University, Aarhus, Denmark, ^14^Institute of Psychiatry, Psychology and Neuroscience, King’s College London, London, UK, ^15^Department of Mental Disorders, Norwegian Institute of Public Health, Oslo, Norway, ^16^Nic Waals Institute, Lovisenberg Diaconal Hospital, Oslo, Norway, ^17^Department of Psychiatry, Universidade Federal do Rio Grande do Sul, Porto Alegre, Brazil, ^18^Adult ADHD Outpatient Program (ProDAH), Hospital de Clínicas de Porto Alegre, Porto Alegre, Brazil, ^19^Section on Negative Affect and Social Processes, Hospital de Clínicas de Porto Alegre, Porto Alegre, Brazil, ^20^School of Medical Sciences, Örebro University, Örebro, Sweden, ^21^Department of Medical Epidemiology and Biostatistics, Karolinska Institutet, Stockholm, Sweden, ^22^Department of Cognitive Neuroscience, Donders Institute for Brain, Cognition and Behavior, Nijmegen, Netherlands, ^23^Department of Biomedicine, University of Bergen, Bergen, Norway, ^24^Division of Psychiatry, Haukeland University Hospital, Bergen, Norway, ^25^Semel Institute for Neuroscience and Human Behavior, UCLA David Geffen School of Medicine, Los Angeles, CA, USA, ^26^Department of Pediatrics, Nemours Children’s Hospital, Wilmington, DE, USA, ^27^Sidney Kimmel Medical College, Thomas Jefferson University, Philadelphia, PA, USA, ^28^Division of Molecular Psychiatry, University of Würzburg, Würzburg, Germany, ^29^Institute of Molecular Medicine, I.M. Sechenov First Moscow State Medical University, Moscow, Russia, ^30^Department of Neuropsychology and Psychiatry, Maastricht University, Maastricht, Netherlands, ^31^Department of Psychiatry, University of California San Diego, La Jolla, CA, USA, ^32^Turner Institute for Brain and Mental Health, Monash University, Melbourne, Australia, ^33^Department of Psychology, The Chinese University of Hong Kong, Hong Kong, China, ^34^Department of Psychiatry, Federal University of São Paulo, São Paulo, Brazil, ^35^Department of Child and Adolescent Psychiatry, Mental Health Services of the Capital Region, Copenhagen, Denmark, ^36^Institute of Clinical Medicine, University of Copenhagen, Copenhagen, Denmark, ^37^National Centre for Register-based Research, Aarhus University, Aarhus, Denmark, ^38^Psychiatric Genetics, QIMR Berghofer Medical Research Institute, Brisbane, Australia, ^39^Departments of Psychiatry and Neuroscience and Physiology, SUNY Upstate Medical University, Syracuse, NY, USA, ^40^Norwegian Institute of Public Health, Oslo, Norway, ^41^Institute of Clinical Medicine, University of Oslo, Oslo, Norway, ^42^Department of Child and Adolescent Psychiatry, Heidelberg University, Heidelberg, Germany

