## Supplementary Figures, Tables and Note for "Statistical and functional convergence of common and rare variant risk for autism spectrum disorders at chromosome 16p"

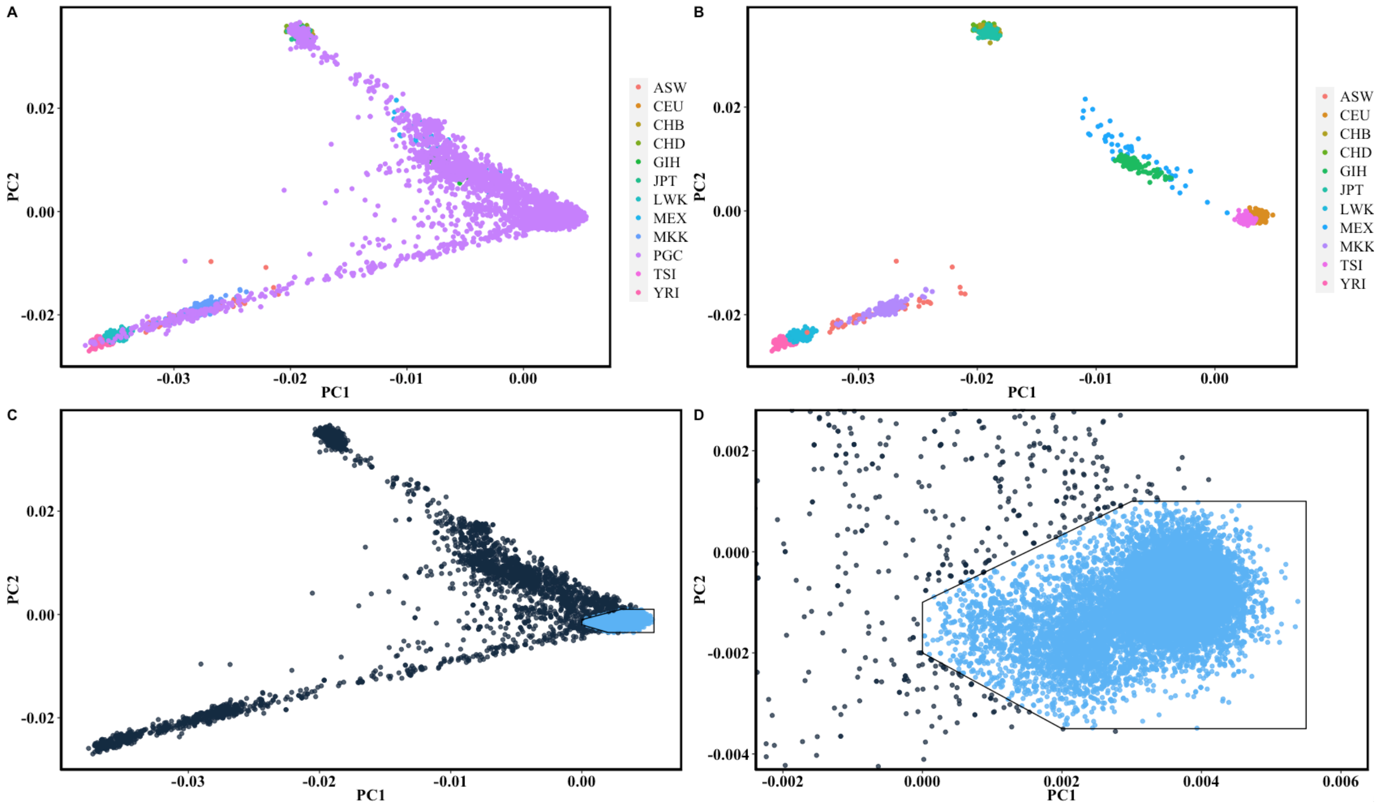

**Supplementary Figure 1: Ancestry analysis of PGC ASD trios**

**(A-B)** The ASD trios from the Psychiatric Genomics Consortium Autism group (PGC) are as described previously (Methods) with the exception of the inclusion of all siblings from multiplex families. We defined a European ancestry subset of PGC for analysis by generating principal components of ancestry using PLINK and by visual inspection relative to Hapmap reference populations. The figure legend lists Hapmap subpopulations and PGC samples. **(C-D)** We defined a family as European ancestry if both parents and proband were European ancestry by PC analysis (4,335 of 5,283 trios, 82%).

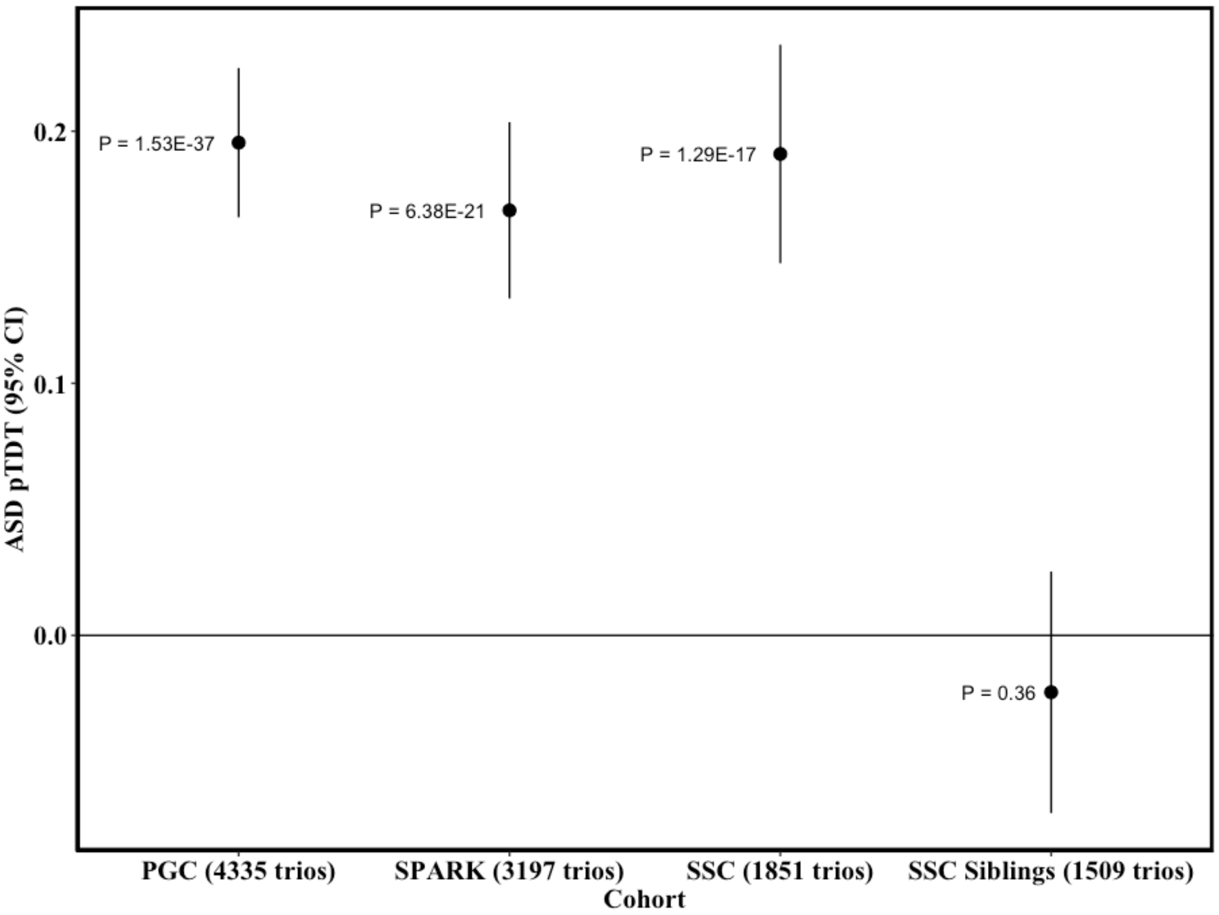

**Supplementary Figure 2: pTDT of ASD PRS in three European-ancestry ASD trio cohorts**

We performed whole genome ASD PRS pTDT using three independent ASD trio datasets as previously described (see Methods). Point estimates are transmission of ASD PRS in units of standard deviations of the mid-parent PRS distribution. Error bars are 95% confidence intervals for transmission. P-values are from two-sided one-sample t-tests of the null that transmission equals 0.

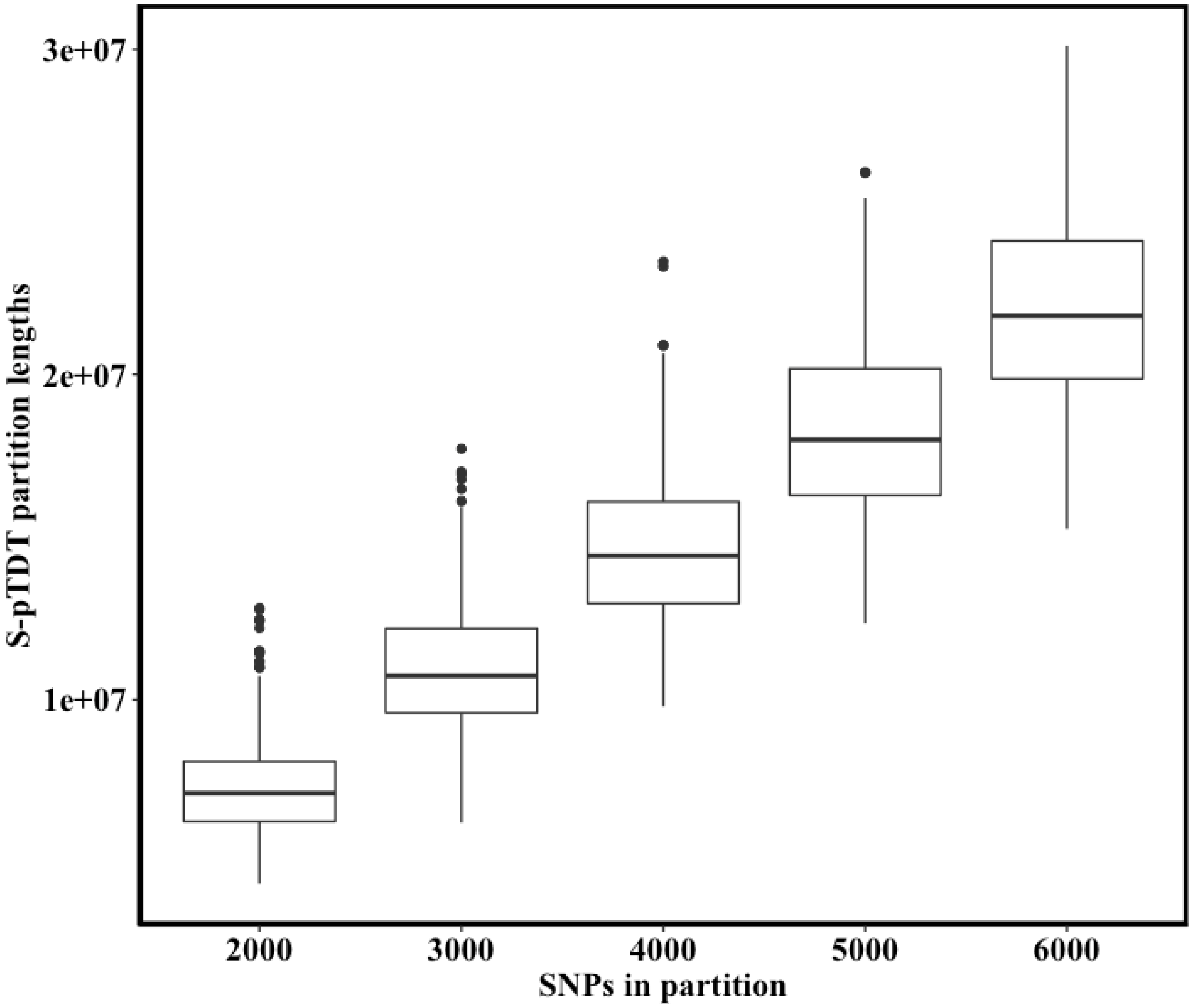

**Supplementary Figure 3: S-pTDT partition lengths in base pairs as a function of number of SNPs in partition**

The distribution of partition lengths in base pairs as a function of number of SNPs in the partition. If the distance between consecutive SNPs is greater than 1Mb, the counted distance is reduced to 1Mb (Methods). We plot here both forward and reverse partitions (i.e. starting at the beginning and end of chromosomes) of count 2,000 SNPs (n = 704), 3,000 SNPs (n = 464), 4,000 SNPs (n = 346), 5,000 SNPs (n = 272), and 6,000 SNPs (n = 220).

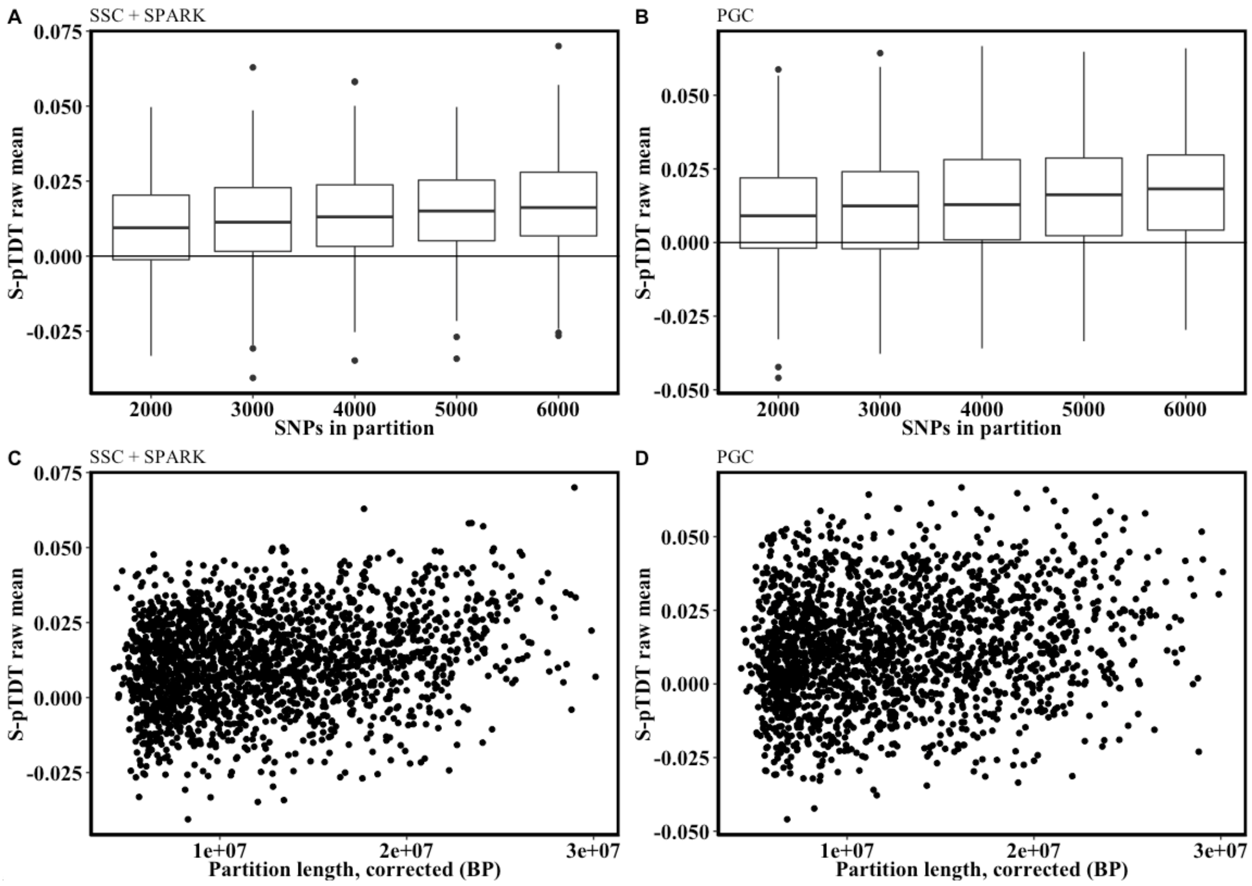

**Supplementary Figure 4: Mean ASD S-pTDT as a function of number of SNPs in partition and partition base pair size in SSC+SPARK and PGC ASD trios**

The relationship between number of SNPs in partition, corrected partition length, and S-pTDT transmission. **(A-B)** S-pTDT increases with the number of SNPs in partition in analysis of the SSC + SPARK cohorts and the PGC trios. **(C-D)** S-pTDT increases with corrected partition length in the SSC + SPARK cohort and the PGC cohort. S-pTDT in units of standard deviations of the mid-parent PRS distribution.

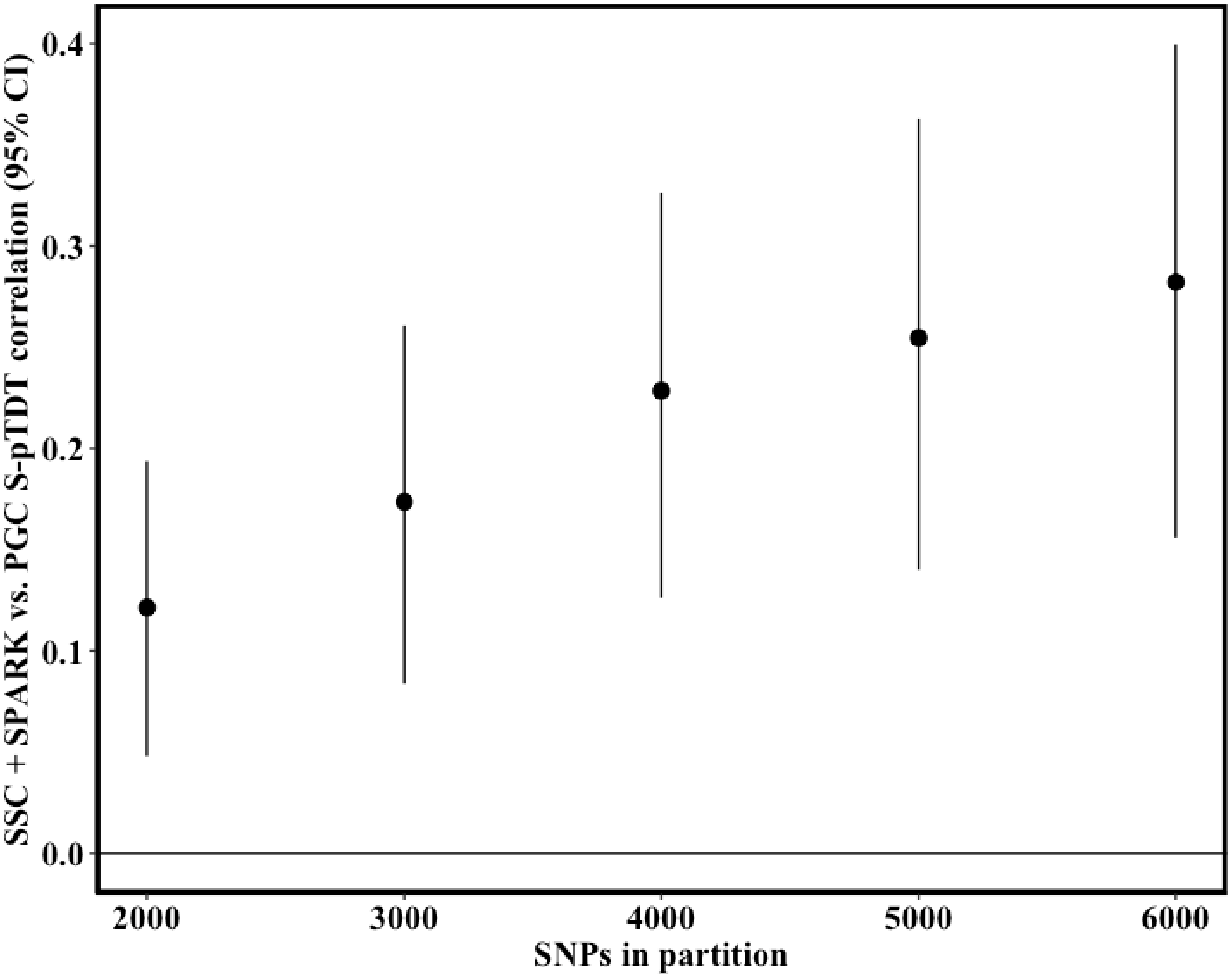

**Supplementary Figure 5: S-pTDT correlation between SSC/SPARK and PGC trios across partition sizes.**

The correlation between S-pTDT values in SSC+SPARK and in PGC. The correlation analysis includes both forward and reverse partitions. As expected, the cross-cohort correlation increases with the number of SNPs in partition (i.e. as more polygenic signal is included in the partition). Error bars are 95% confidence intervals on the mean of the S-pTDT transmission.

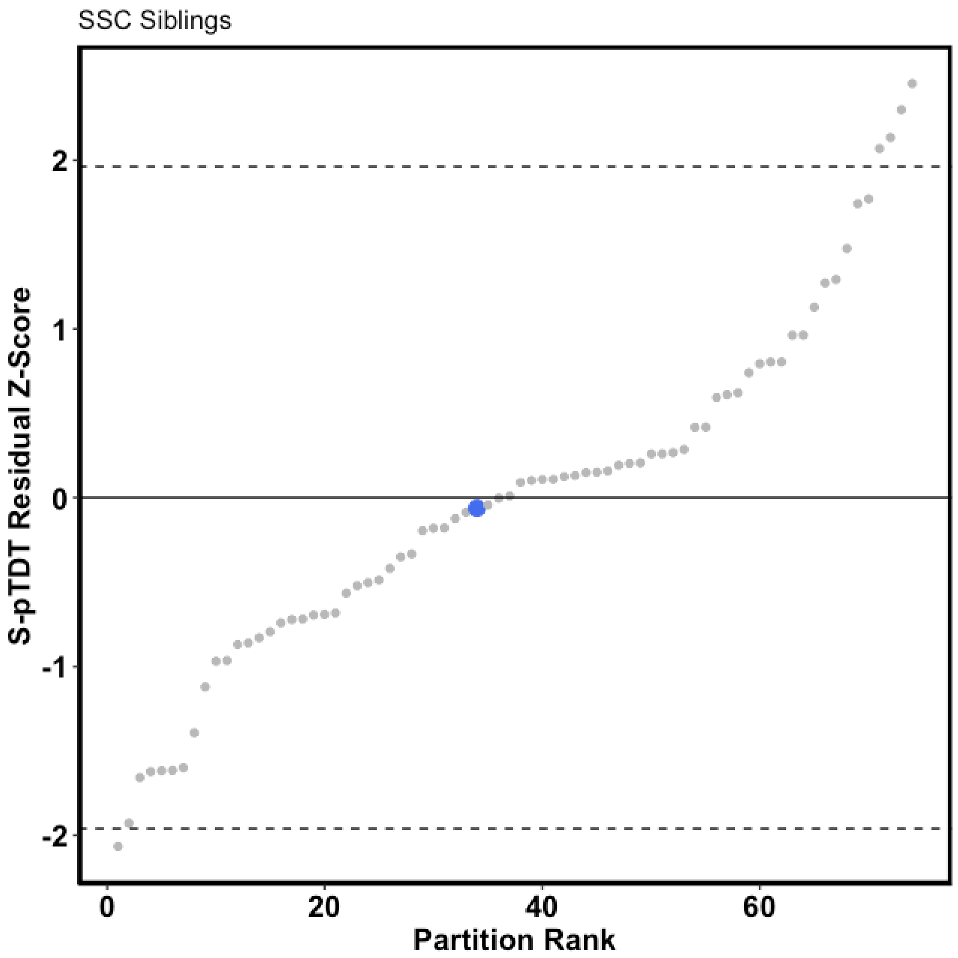

**Supplementary Figure 6: 16p ASD PRS is not over-transmitted to 1,509 unaffected siblings in SSC**. An identical analysis to that presented in Figure 1D, except instead of testing for transmission of 16p PRS from parents to probands, here we test transmission from parents to unaffected siblings. The blue dot denotes the 16p partition, while the other gray dots are the 73 other control partitions. The y-axis is the S-pTDT z-score.

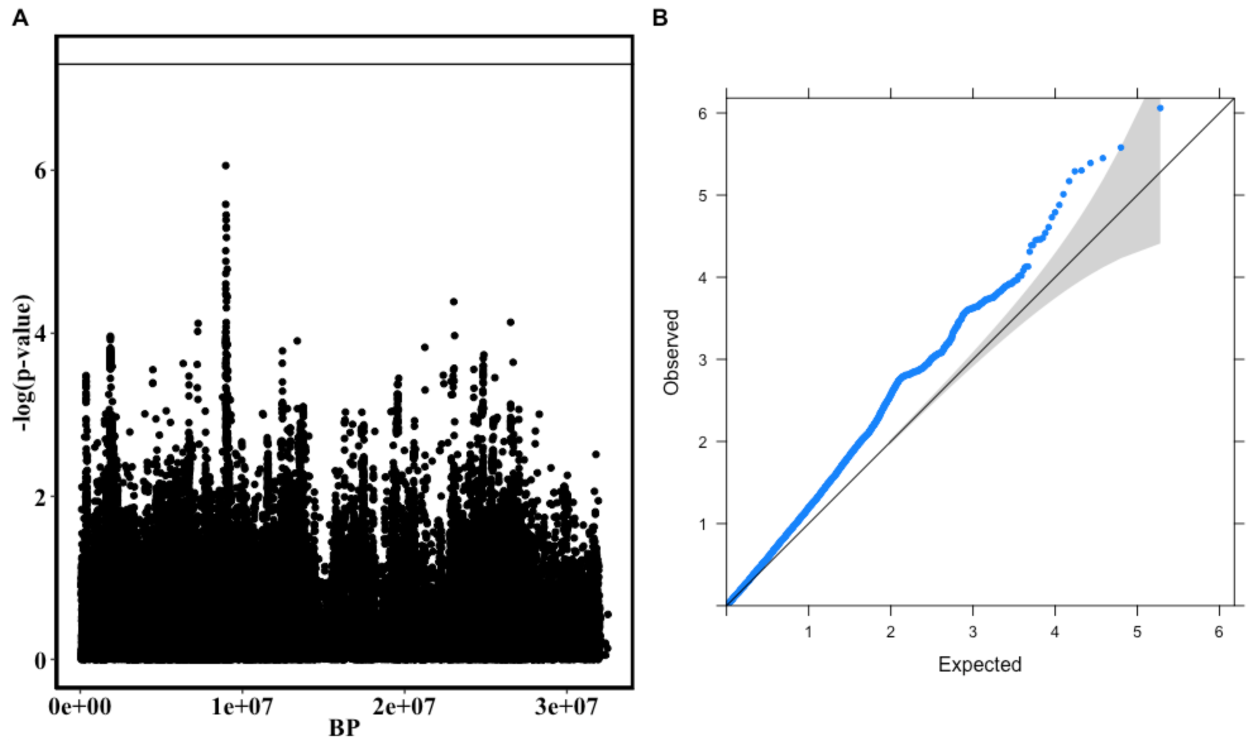

**Supplementary Figure 7: ASD GWAS at 16p**

**(A)** Local manhattan plot visualization of ASD GWAS on 16p. Each point is a SNP, with the x-axis value its position on chromosome 16 and the y-axis value the -log(p-value, base 10) from the GWAS. The GWAS is an unpublished meta-analysis of the iPSYCH and PGC collections (26067 cases, 46455 controls). **(B)** The same SNPs as (A), here represented as a quantile-quantile plot (code adapted from Matthew Flickinger, https://genome.sph.umich.edu/wiki/Code_Sample:_Generating_QQ_Plots_in_R). Confidence interval represents alpha = 0.05.

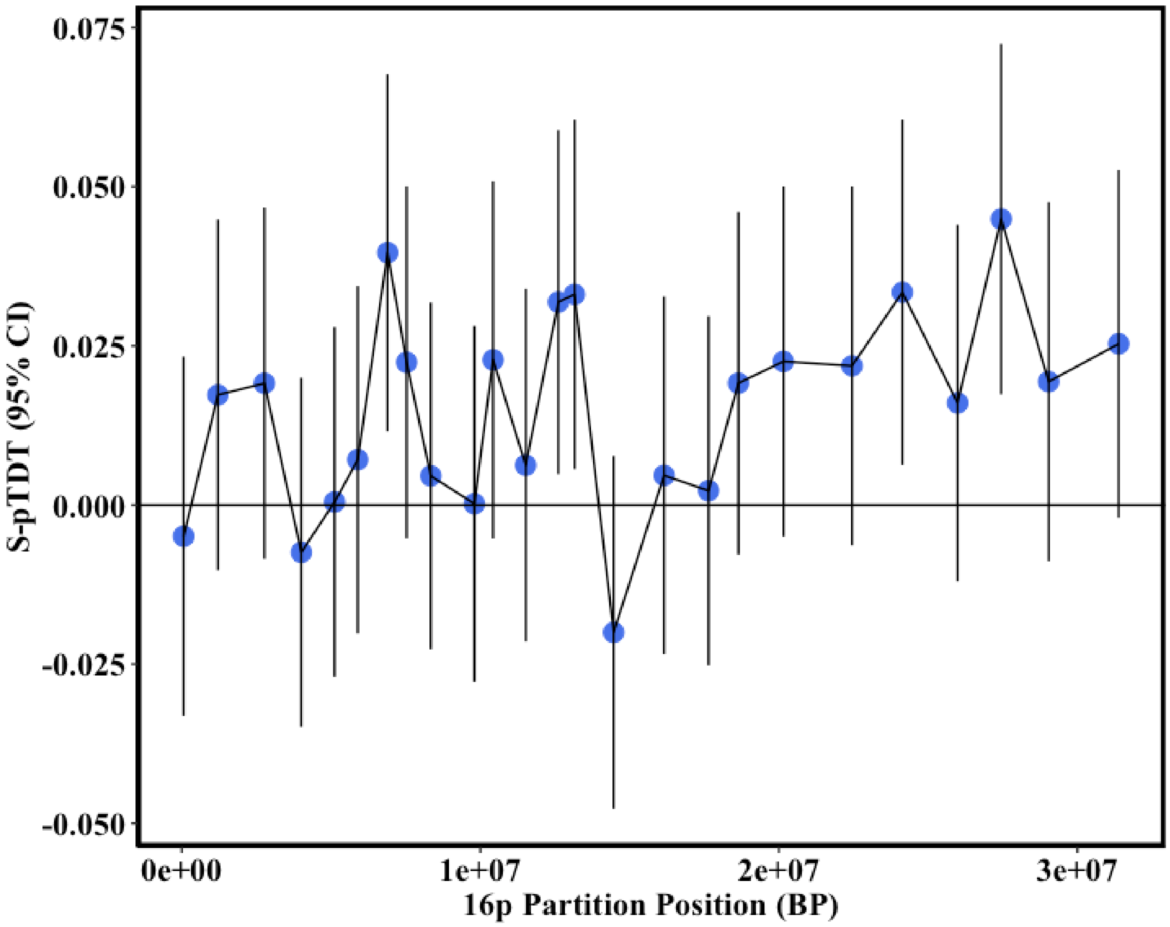

**Supplementary Figure 8: ASD S-pTDT in SSC + SPARK in LD-independent blocks at 16p**

We performed S-pTDT in 25 LD-independent blocks on chromosome 16p using the SSC + SPARK trios and the iPSYCH-only ASD GWAS. The LD-independent blocks are from a previous publication (Methods). The blue dots are the mean S-pTDT value in SSC + SPARK using an ASD PRS constructed from SNPs in that block, in units of SDs of the mid-parent PRS distribution. The x-axis position of the blue dots is the midpoint of the block. The error bars are 95% CI of the S-pTDT mean.

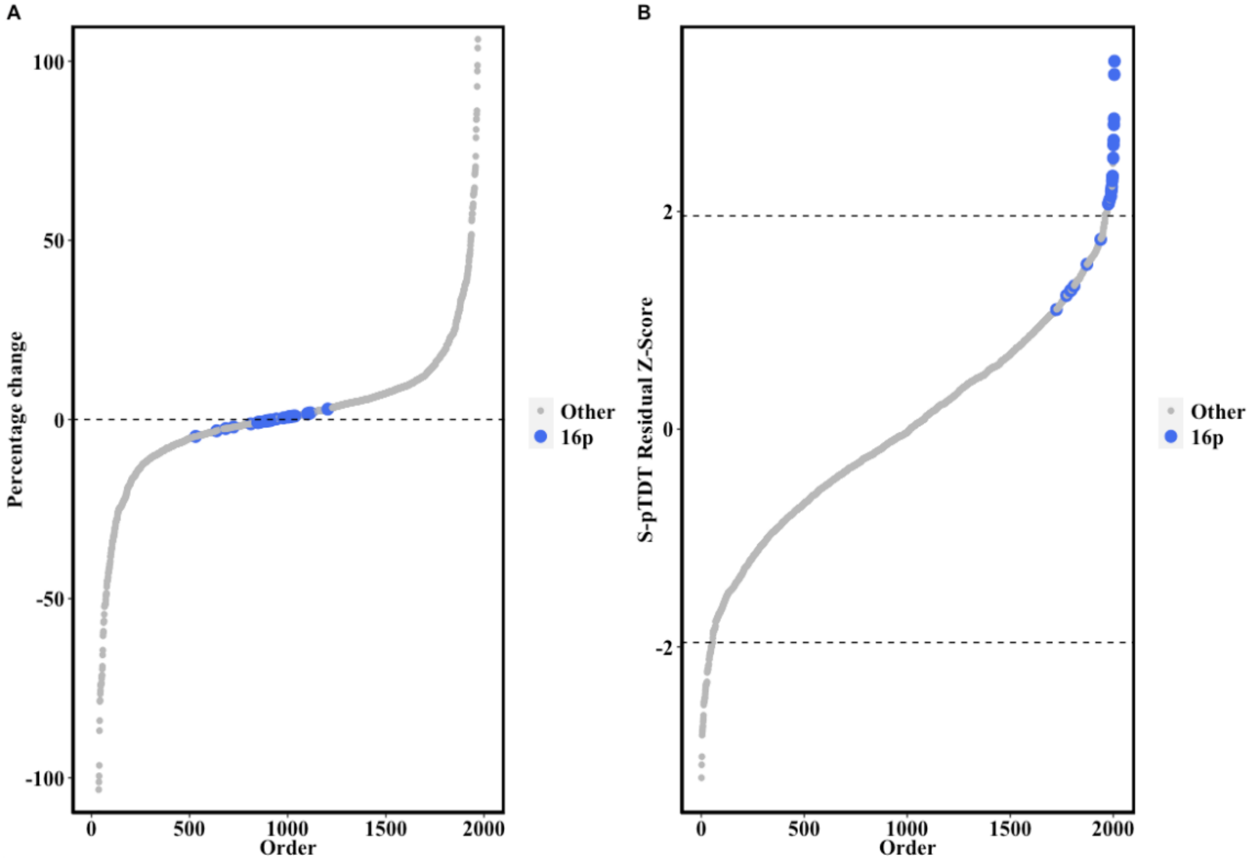

**Supplementary Figure 9: ASD S-pTDT without SSC/SPARK probands carrying 16p CNV**

We tested the hypothesis that ASD probands with a neurodevelopmental-disorder associated CNV on 16p in SSC + SPARK drive the S-pTDT signal by identifying these probands and removing them from analysis. We defined genomic-disorder associated CNV based existing literature curation (Methods). Of the 5,048 trios in the SSC + SPARK analysis, we removed 51 (1.0%) where a proband carried at least one of this class of CNV (either inherited or *de novo*) on the p-arm of chromosome 16. **(A)** S-pTDT estimates do not qualitatively change after removal of these 51 trios, with small percentage change in S-pTDT estimates for the 16p partitions (blue) relative to the change in other partitions (gray). **(B)** We also find that the 16p partitions remain with large residual z-scores after removal of these trios.

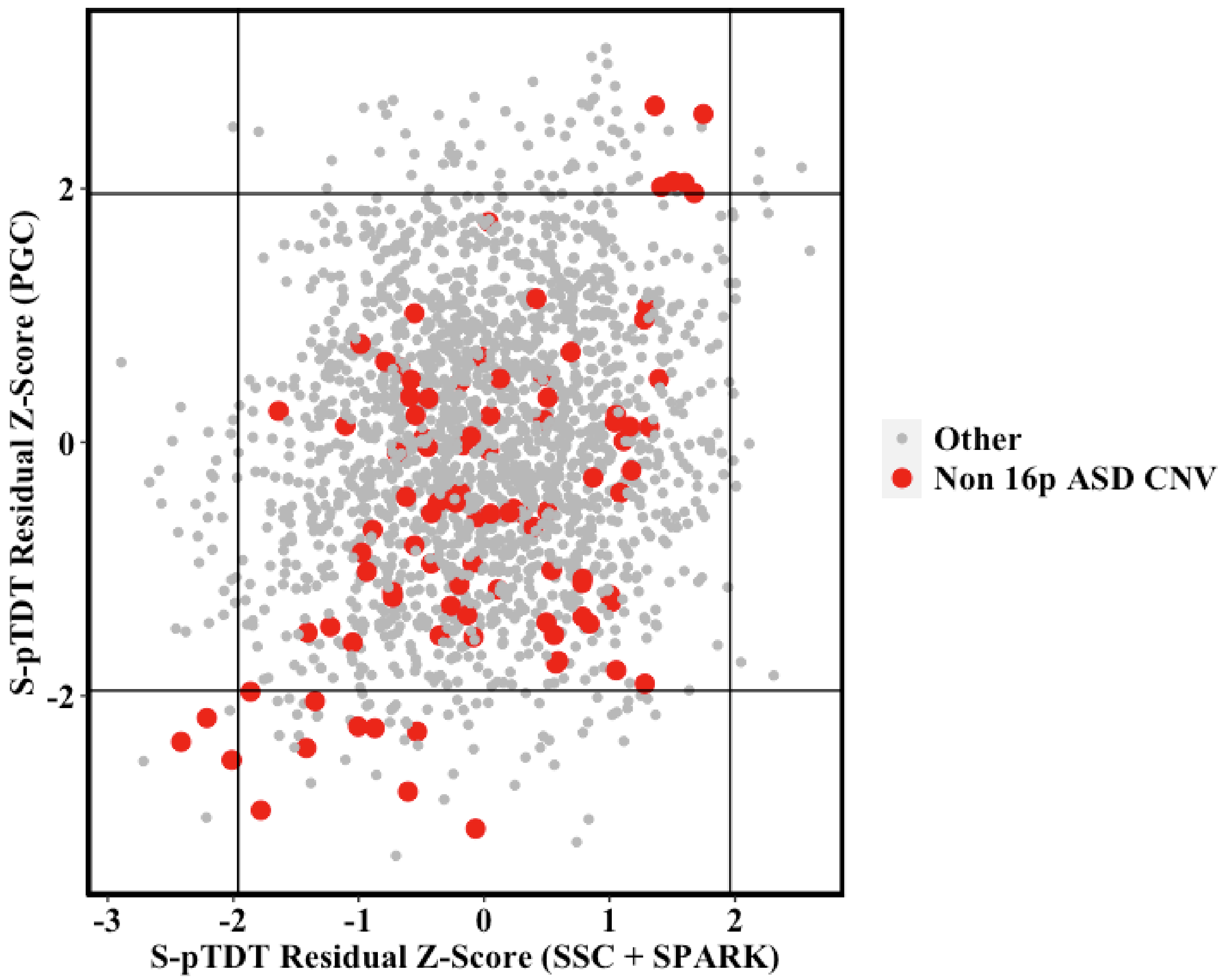

**Supplementary Figure 10: ASD S-pTDT highlighting partitions including ASD-associated CNV**

Primary ASD S-pTDT result (Figure 1B), highlighting partitions in red that include an ASD-associated CNV. We defined ASD-associated CNV as those listed in SFARI Gene (Supplementary Table 7). The axes are residual z-scores from the S-pTDT model (Methods), for SSC+SPARK (x-axis) and PGC (y-axis).

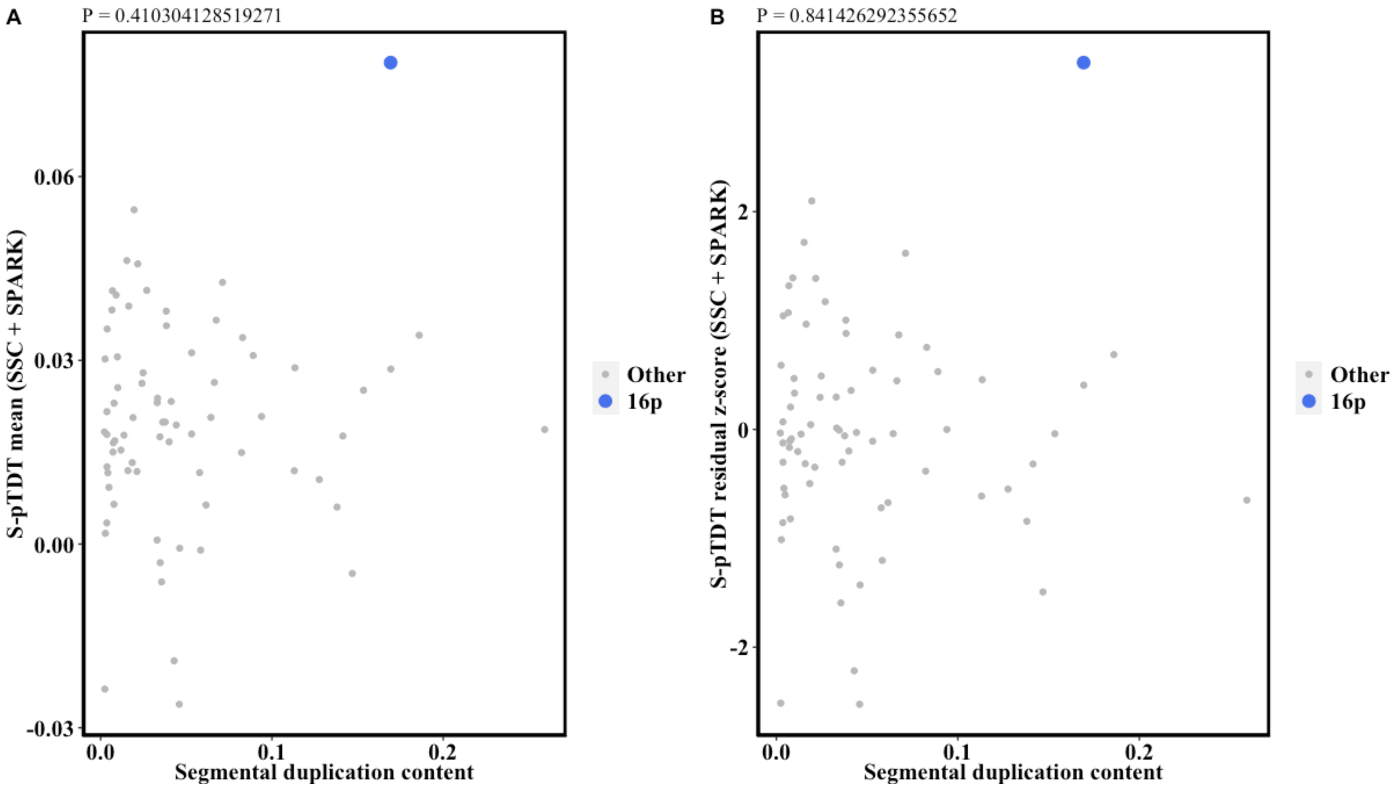

**Supplementary Figure 11: Association between segmental duplication content and ASD S-pTDT**

**(A)** Association between segmental duplication content per 33Mb partition and ASD S-pTDT mean in SSC+SPARK. The p-value at the top of the plot is from the correlation test of segmental duplication content and S-pTDT mean. The 16p partition is noted in blue. **(B)** As in (A), except the y-axis is now the residual z-score from the ASD S-pTDT model (Methods).

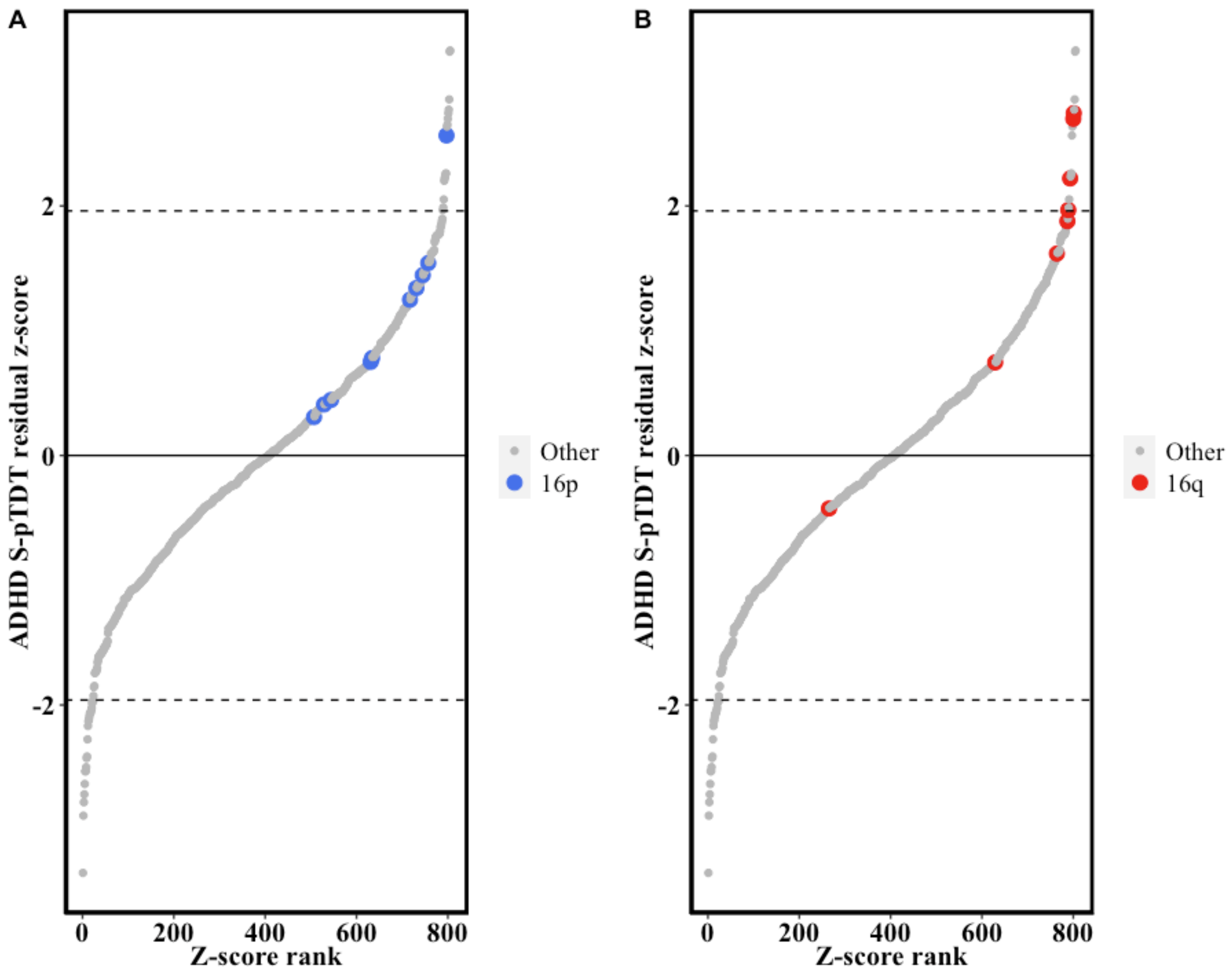

**Supplementary Figure 12: ADHD S-pTDT**

**(A)** We performed S-pTDT using ADHD trios (n = 1,634 European ancestry trios from the PGC) and an ADHD PRS generated from a non-overlapping ADHD GWAS from the iPSYCH collection (25,895 cases, 37,148 controls). The partitions located on 16p are denoted in blue. The 16p partition with marginal significance (5,000 SNPs, start BP: 28,861,734, end BP: 69,367,996, residual z-score: 2.56) has 4,807/5,000 (96%) of SNPs beyond the 16p boundary, suggesting that it reflects signal on 16q. **(B)** This hypothesis is confirmed when we highlight the position of 16q partitions, many of which are marginally significant in 16q.

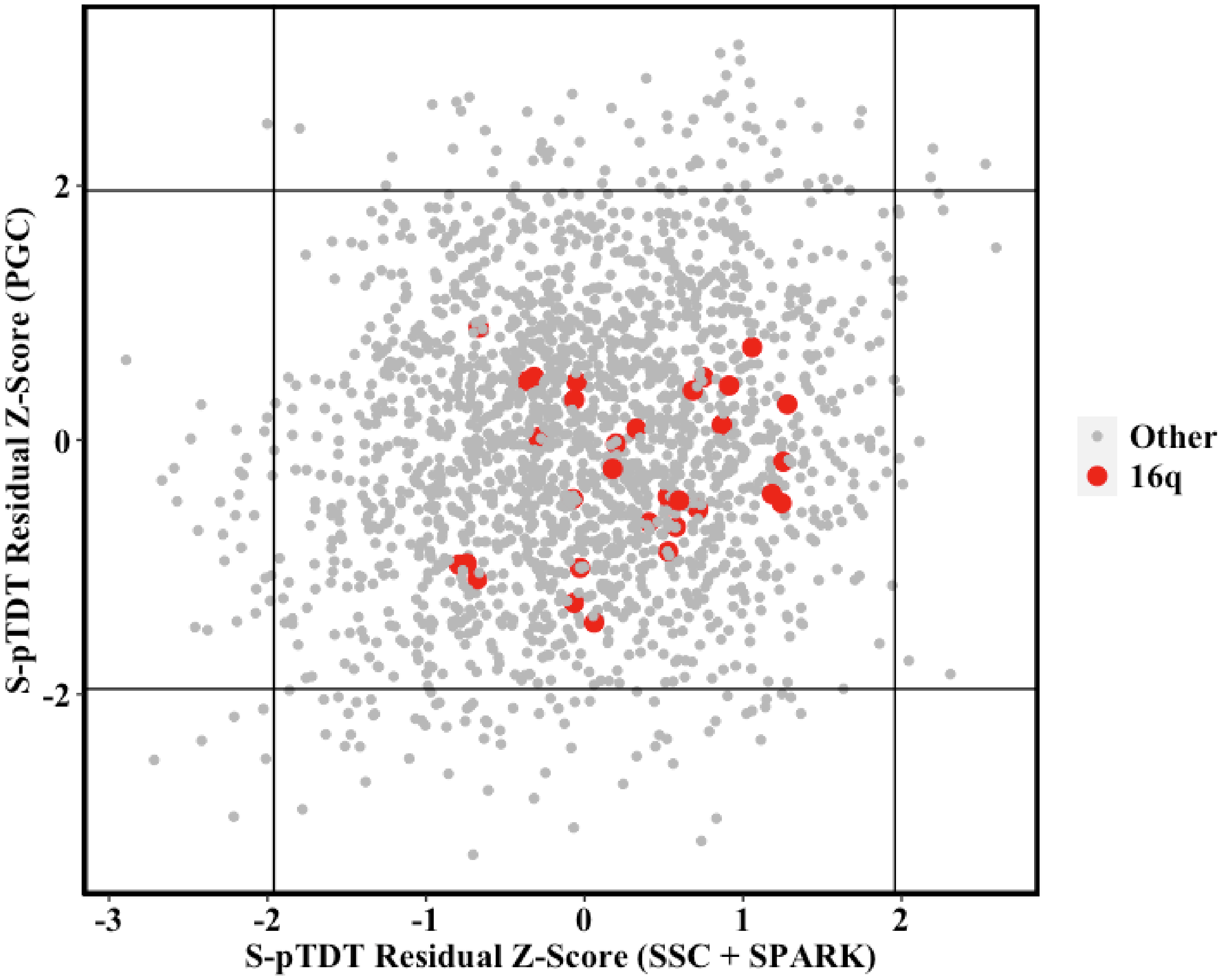

**Supplementary Figure 13: ASD S-pTDT for partition on 16q**

Primary ASD S-pTDT association result (Figure 1B), with partitions in red on chromosome 16 that are not located on the p-arm (start position > 33Mb). As opposed to partitions on the p-arm, partitions on the q-arm are not enriched in high S-pTDT values. The axes are residual z-scores from the S-pTDT model (Methods), for SSC+SPARK (x-axis) and PGC (y-axis).

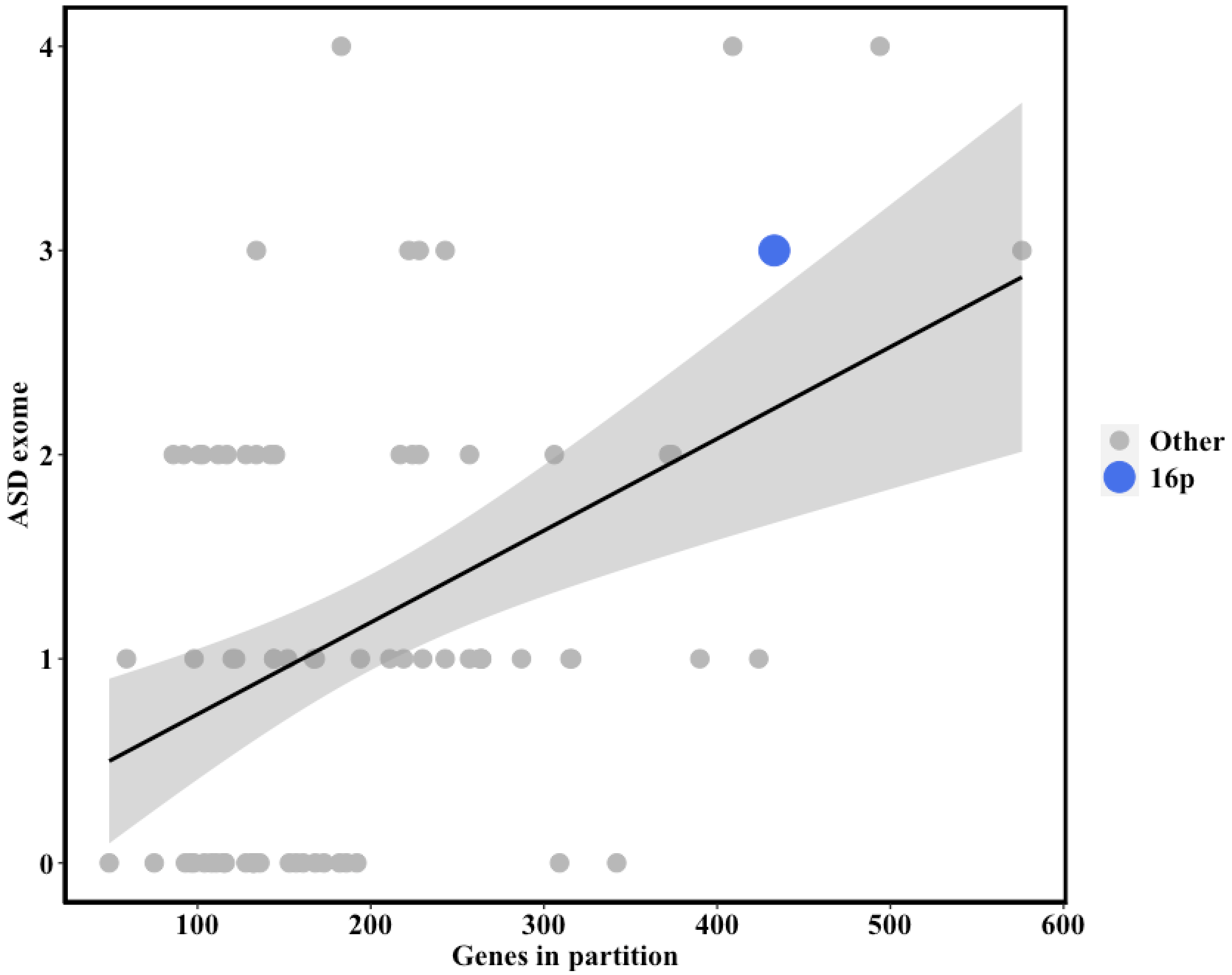

**Supplementary Figure 14: Association between total genes and genes implicated in ASD from exome association studies**

Across 33Mb partitions, the relationship between the number of genes in partition (x-axis) and the number of genes implicated in ASD via rare variant exome-association studies (TADA q-value < 0.1 from Satterstrom 2020) (y-axis). Each point is a 33Mb region, with the 16p region highlighted in blue. The shaded region denotes a 95% CI.

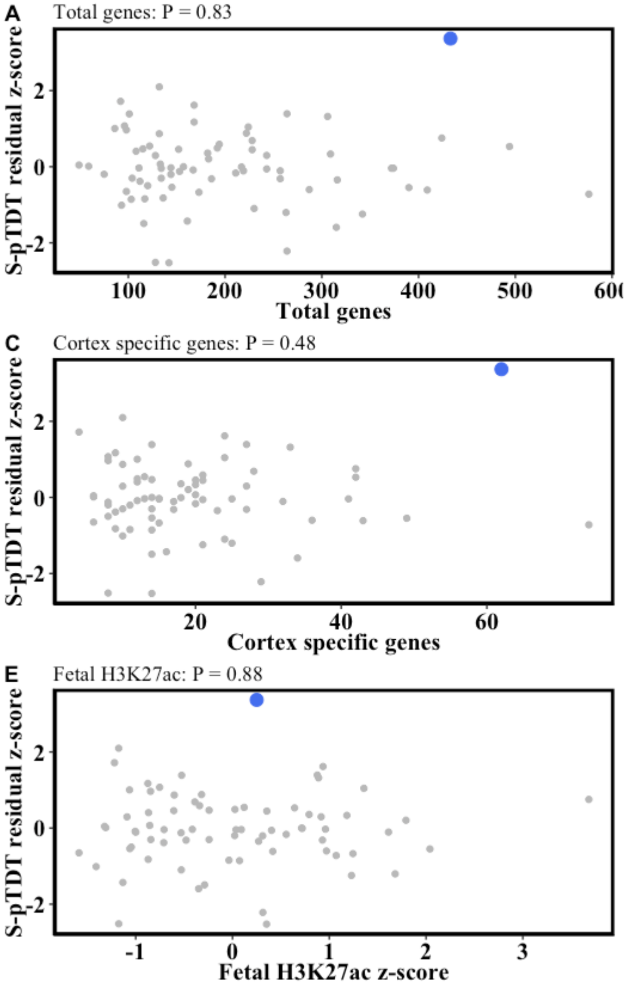

**Supplementary Figure 15: Association between gene/enhancer density and S-pTDT**

No association between ASD S-pTDT signal (residual z-score) and markers of region gene density for the 33Mb partitions. P-values above plots are from the x-y correlation. **(A)** There is no relationship between the number of genes in the partition and normalized S-pTDT signal (P = 0.83). **(B)** There is no relationship between the number of constrained genes (probability of loss-of-function intolerance ≧ 0.9) and normalized S-pTDT signal (P = 0.85). **(C)** There is no relationship between the number of genes specifically expressed in cortex (defined as top 10% highest cortex vs. non-brain tissue t-statistic from Finucane 2018) and S-pTDT signal (P = 0.48). **(D)** There is no relationship between the number of genes in the partition implicated in ASD via rare variant exome-association studies (Methods) and the S-pTDT signal (P = 0.21). **(E)** There is no relationship between the density of accessible fetal brain enhancers (H3K27ac) (z-score normalized) and S-pTDT signal (P = 0.88).

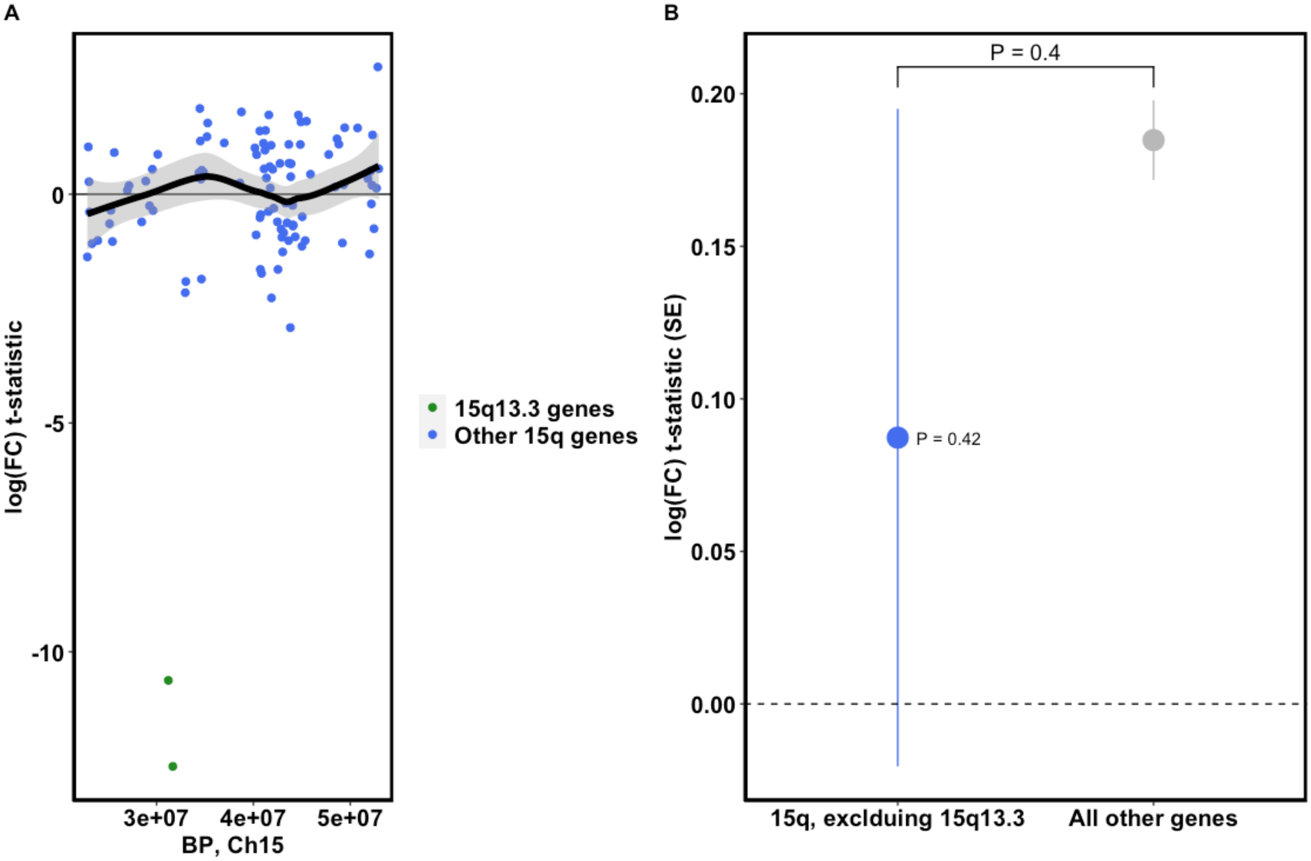

**Supplementary Figure 16: *In vitro* deletion analysis of 15q13.3**. Induced pluripotent stem cells undergo CRISPR-Cas9-mediated deletion of the 15q13.3 CNV region, differentiation into induced neurons and transcriptome profiling with RNA-seq (n = 11 biological replicates). Differential expression analysis compares these samples to controls (n = 6 biological replicates) without deletion of the CNV. **(A)** Differential expression of 15q brain genes after deletion of 15q13.3 CNV. Brain genes are defined based on above median normalized expression level of genes over all samples in analysis. Genes in the deletion region +/- 0.1Mb are green, while all other genes on 15q are in blue. The y-axis is the $\log_2$(fold-change) t-statistic per gene. The trend line is for genes outside of the CNV (blue dots) with a 95% CI shaded in gray. **(B)** 15q13.3 CNV effect on 15q genes is not different from its effect on all other genes. Point estimates are of mean differential expression t-statistic for the group of genes +/- SE. Top p-value is from two-sample t-test comparing groups.

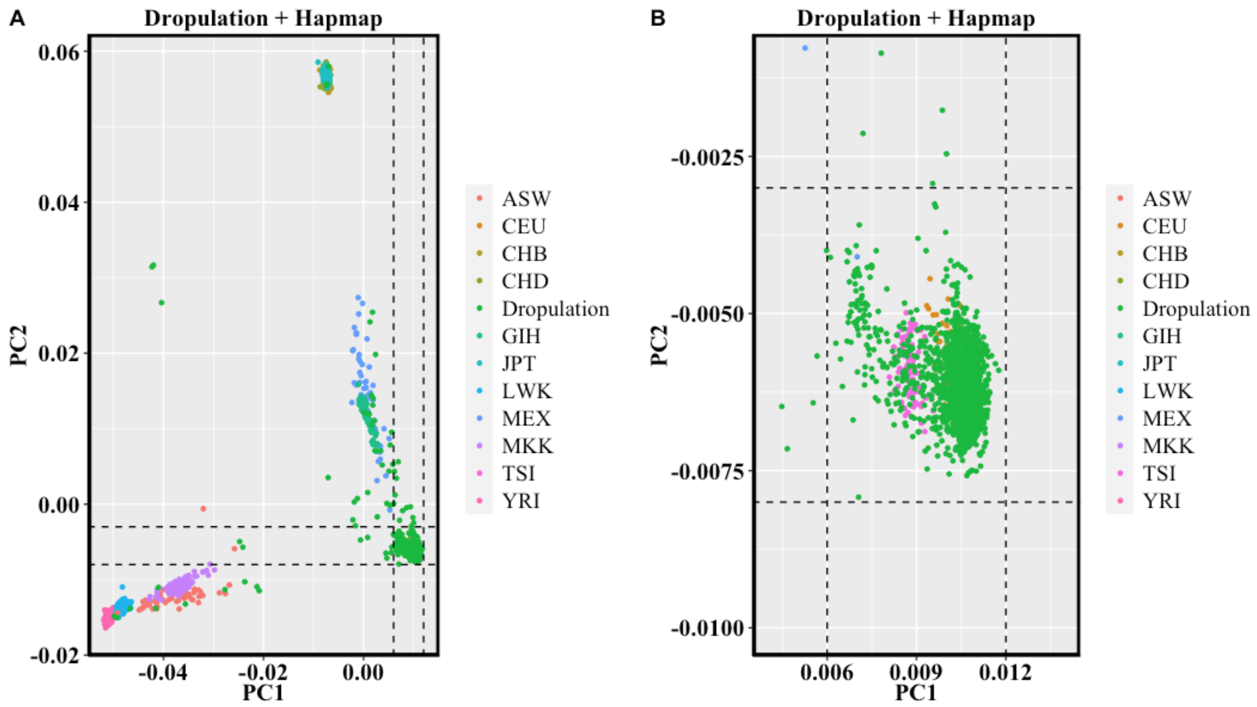

**Supplementary Figure 17: Ancestry analysis of single-nucleus RNA-seq donor samples**

We performed principal components analysis to identify ancestrally homogeneous subgroups of the single-nucleus RNA-seq donor samples. Since the majority of the samples were of European ancestry by visual inspection of the PCs, we defined a European ancestry subgroup for analysis based on alignment with European ancestry Hapmap samples. We removed regions of long-range LD before computing principal components with PLINK. **(A)** All Hapmap and Dropulation samples in principal component space, colored by group. Abbreviations in the legend correspond to Hapmap subgroups. Dropulation samples in the dashed square are defined as European ancestry and retained for downstream analysis (1707/1770 samples, 96%) **(B)** Same as (A), magnified.

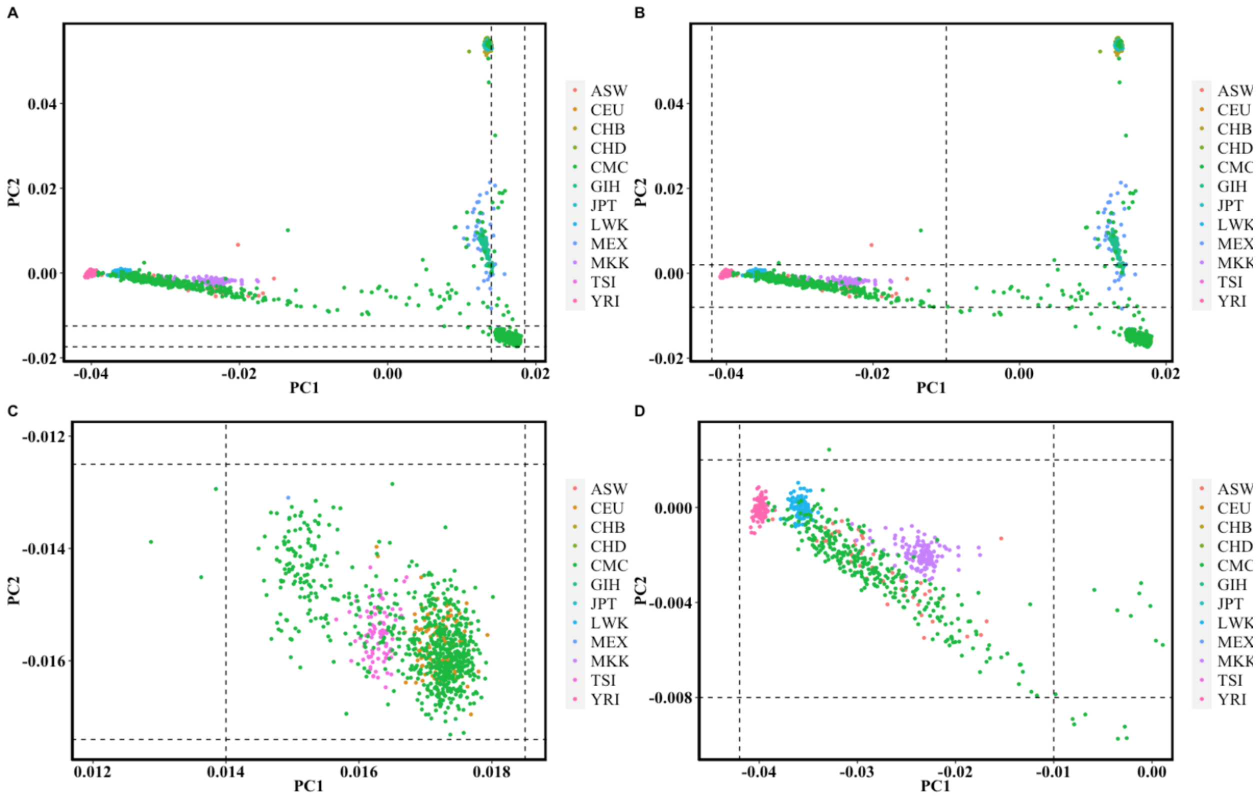

**Supplementary Figure 18: Ancestry analysis of CommonMind samples**

We performed principal components analysis to identify ancestrally homogeneous subgroups of the CommonMind samples. We merged the CommonMind samples with Hapmap samples for ancestral reference and removed regions of long-range LD before calculating principal components of ancestry using PLINK. **(A,C)** We defined a European ancestry subgroup of the CommonMind samples by visual inspection of alignment with Hapmap samples; we retained samples within the dashed boundaries (694/1,076 of the genotyped samples, 64%). **(B,D)** We defined an African ancestry subgroup of the CommonMind samples by visual inspection of alignment with Hapmap samples; we retained samples within the dashed boundaries (299/1,076 of the genotyped samples, 28%).

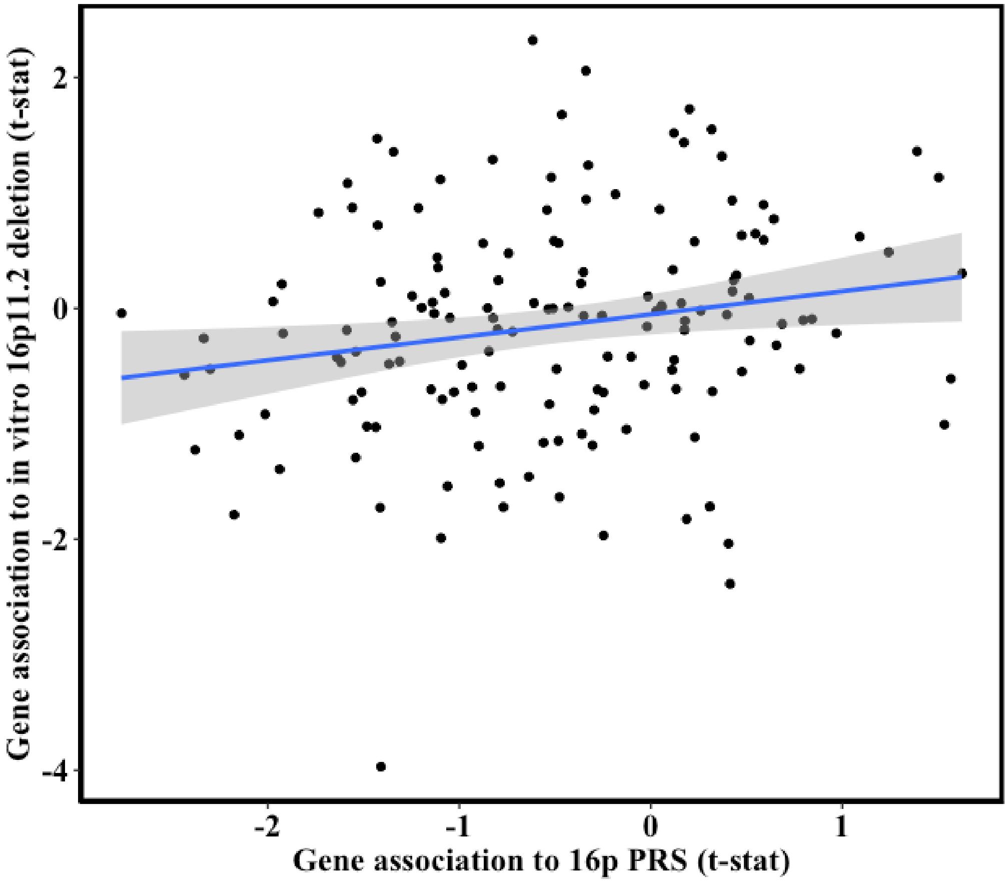

**Supplementary Figure 19: Correlation between gene association to 16p PRS and to 16p11.2 *in vitro* deletion**

The positive correlation of 16p genes in their association to 16p PRS and to the 16p11.2 *in vitro* deletion. The x-axis shows the association t-statistics from the all sample meta-analysis of 16p PRS and 16p gene expression. The y-axis shows the association t-statistics from the 16p11.2 *in vitro* deletion analysis. The shaded region is 95% CI. This plot is the same as Figure 4A with the inclusion of the outlying point.

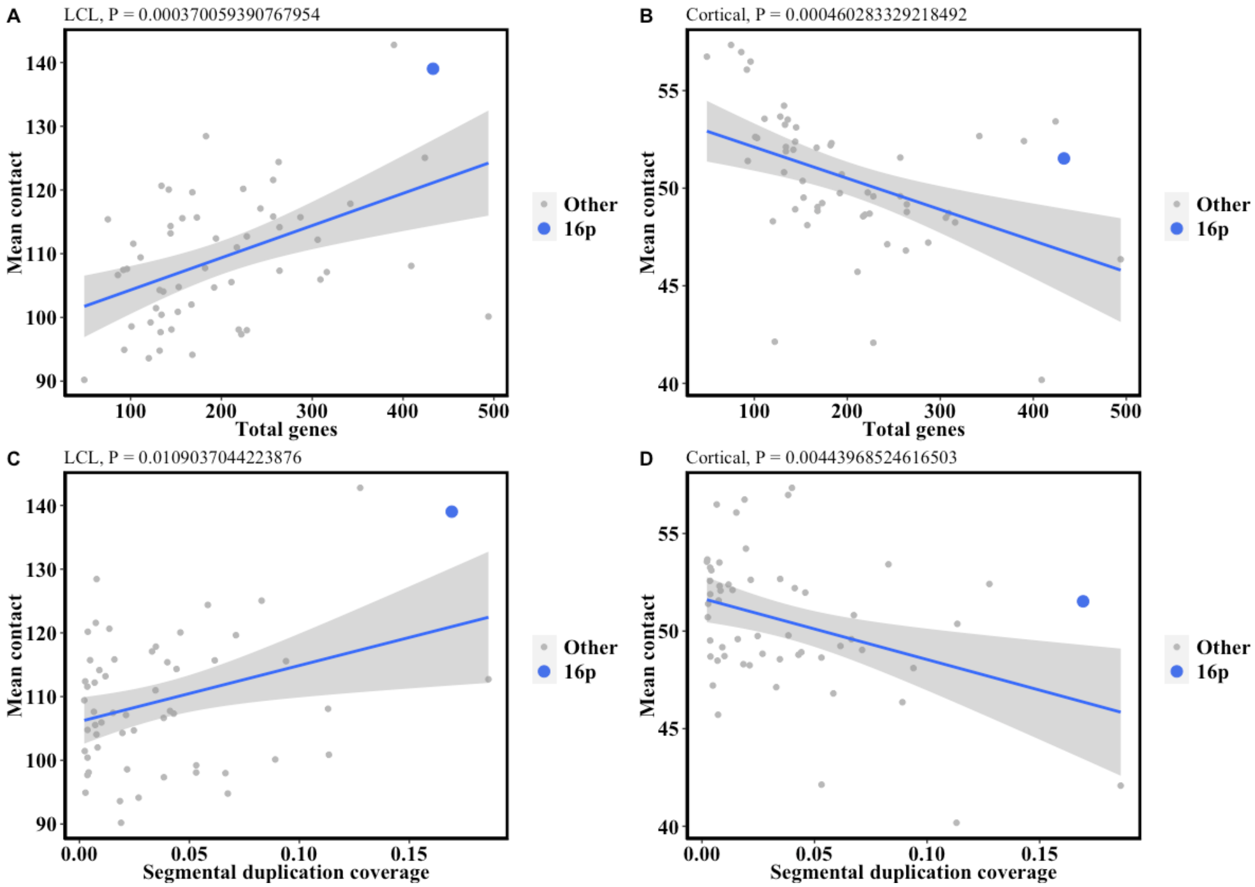

**Supplementary Figure 20: Association between segmental duplication content / gene density and Hi-C mean content**

The relationship between gene density, segmental duplication content and mean contact frequency in two Hi-C datasets, one of lymphoblastoid cell lines and one of mid-gestational cortical plate (Methods). Each point is a 33Mb partition (see Methods). Mean contact is the mean within-partition off-diagonal value from the contact matrix. Segmental duplication coverage is defined in main text Methods. The above-plot p-value is from the correlation test between the x and y variables as shown.

**Supplementary Note 1: Derivation of pTDT for case-pseudocontrol genotypes**

The genotypes for PGC trios are imputed as cases and pseudocontrols, where pseudocontrol genotypes are constructed from the alleles that are not transmitted from parents to offspring. Below we derive a pTDT estimator using cases and pseudocontrols, instead of the traditional input of cases and both parental genotypes.

pTDT is defined as the standardized difference between case PRS and the expectation defined by the mid-parent PRS:

$$pTDT = \frac{PRS_{case} - PRS_{MP}}{SD(PRS_{MP})}$$

The union of the genotypes of the case and the pseudocontrol constitute the transmitted and untransmitted alleles from the parents. Thus, the union of the case and pseudocontrol genotypes, divided by 2, equals the average parental genotype, and by extension, the mid-parent PRS:

$$PRS_{MP} = \frac{PRS_{case} + PRS_{pseudocontrol}}{2}$$

Thus, by substitution of the expression for $PRS_{MP}$ into the pTDT equation above, we derive an estimator compatible with case-pseudocontrol genotypes.

| Name | Cases | Controls | Total samples | SNPs after LDpred | Reference |
|---|---|---|---|---|---|
| iPSYCH ASD | 19870 | 39078 | 58948 | 1152500 | iPSYCH consortium |
| iPSYCH+PGC ASD | 26067 | 46455 | 72522 | 1169771 | iPSYCH, PGC consortia |
| iPSYCH ADHD | 25895 | 37148 | 63043 | 1090116 | iPSYCH consortium |

**Supplementary Table 1: GWAS summary statistics used in analysis**

| **ASD family cohorts (European ancestry)** | ***Complete families*** | ***Unique families*** | **Access** |
|---|---|---|---|
| SSC (parents and proband) | 1851 | 1851 | SFARI |
| SSC (parents and unaffected sibling) | 1509 | 1509 | SFARI |
| SPARK (multiplex families, parent cases removed) | 3197 | 2880 (2596 with 1 proband, 253 with 2, 29 with 3, 2 with 4) | SFARI |
| Psychiatric Genomics Consortium (multiplex families) | 4335 | 3258 (2312 with 1 proband, 829 with 2 probands, 104 with 3, 12 with 4, 1 with 5) | PGC |
| **ADHD family cohorts (European ancestry)** | ***Complete families*** | ***Unique families*** | **Access** |
| Psychiatric Genomics Consortium (parents and proband) | 1634 | 1634 | PGC |
| **Genotyped-expression cohorts** | **Total n** | **Schizophrenia diagnosis (n)** | **Access** |
| Dropulation donors, European ancestry | 122 | 58 | McCarroll Lab |
| Commonmind, African ancestry | 194 | 67 | CommonMind/Synapse |
| Commonmind, European ancestry | 238 | 68 | CommonMind/Synapse |

**Supplementary Table 2: Cohorts used in analysis**
